# Rapid and lasting generation of B-cell memory to SARS-CoV-2 spike and nucleocapsid proteins in COVID-19 disease and convalescence

**DOI:** 10.1101/2020.11.17.20233544

**Authors:** Gemma E. Hartley, Emily S.J. Edwards, Pei M. Aui, Nirupama Varese, Stephanie Stojanovic, James McMahon, Anton Y. Peleg, Irene Boo, Heidi E. Drummer, P. Mark Hogarth, Robyn E. O’Hehir, Menno C. van Zelm

## Abstract

**Background:** Lasting immunity to SARS-CoV-2 following infection is questioned because serum antibodies decline in convalescence. However, functional immunity is mediated by long-lived memory T and B (Bmem) cells.

**Objective:** To determine the longevity and immunophenotype of SARS-CoV-2-specific Bmem cells in COVID-19 patients.

**Methods:** Recombinant spike receptor binding domain (RBD) and nucleocapsid protein (NCP) were produced for ELISA-based serology, and biotinylated for fluorescent tetramer generation to identify SARS-CoV-2-specific Bmem cells by flow cytometry with a panel of 13 mAbs. 36 blood samples were obtained from 25 COVID-19 patients (11 paired) between 4-242 days post-symptom onset for detection of neutralizing antibodies, IgG serology and flow cytometry.

**Results:** The recombinant RBD and NCP were specifically recognized by serum IgG in all patients and reactivity declined >20 days post-symptom onset. All patients had detectable RBD- and NCP-specific Bmem cells at 8.23-267.6 cells/ml of blood (0.004-0.13% of B cells) regardless of sampling time. RBD- and NCP-specific Bmem cells predominantly expressed IgM or IgG1, with the latter formed slightly later than the former. RBD-specific IgG^+^ Bmem were predominantly CD27^+^, and numbers significantly correlated with circulating follicular helper T cell numbers.

**Conclusion:** RBD- and NCP-specific Bmem cells persisted for 8 months, indicating that the decline in serum antibodies after 1 month does not indicate waning of immunity but a contraction of the immune response. Flowcytometric detection of SARS-CoV-2-specific Bmem cells enables detection of long-term functional immunity following infection or vaccination for COVID-19.

## INTRODUCTION

Coronavirus disease (COVID)-19 is a global health emergency. The causative agent, severe acute respiratory syndrome coronavirus-2 (SARS-CoV-2) is highly contagious and has infected tens of millions worldwide and caused over 1.2 million deaths since its discovery in Wuhan, China in December 2019 (Richardson et al., 2020; Williamson et al., 2020). Although mild or asymptomatic in many cases, SARS-CoV-2 infection in the elderly and individuals with chronic health problems can result in severe COVID-19 requiring invasive ventilation or in death (Meyts et al., 2020; Ruan et al., 2020; Wang et al., 2020; Yang et al., 2020).

Since early 2020, many insights have been obtained into the pathology of severe COVID-19. It appears that high viral loads induce strong inflammatory responses that cause systemic disease, especially in the elderly and in individuals with co-morbidities requiring immunosuppression (Ruan et al., 2020). Immunomodulation with corticosteroids has improved survival in hospitalized individuals, and anti-SARS-CoV-2 monoclonal antibody treatments have shown early evidence of alleviating symptoms and decreasing SARS-CoV-2 viral load in mild disease (Beigel et al., 2020; Chen et al., 2020).

The COVID-19 pandemic has led to a huge global effort to identify a safe therapeutic vaccine to induce protective immunity. Our current understanding of SARS-CoV-2 immunity is based mainly on previous experiences with SARS-CoV, supplemented with recent studies in patients infected with and recovered from SARS-CoV-2 infection. Similar to SARS-CoV infection (Huang et al., 2004; Leung et al., 2004), the main antibody targets in SARS-CoV-2 are the spike and nucleocapsid proteins (NCP) (Burbelo et al., 2020; Grzelak et al., 2020; Long et al., 2020; Wilson et al., 2020). These antibodies are detectable from approximately 6 days after PCR confirmation of infection, and those directed against spike receptor binding domain (RBD) show neutralizing capacity and hence, can prevent infection (Seydoux et al., 2020; Suthar et al., 2020). However, the rapid decline of anti-SARS-COV-2 serum IgG levels beyond 20 days post-diagnosis and the transient presence of circulating plasmablasts have led to questions about the longevity of immunity (Gudbjartsson et al., 2020; Ibarrondo et al., 2020; Newell et al., 2020; Ng et al., 2020; Patel et al., 2020). In contrast, antigen-specific memory T cells and memory B (Bmem) cells can be detected in convalescence (Juno et al., 2020; Sekine et al., 2020). As these memory cells are programmed to respond rapidly upon subsequent antigen encounter, it is reasonable to hypothesize that these long-lived memory cells provide durable long-term immunity (Cox and Brokstad, 2020; Nguyen-Contant et al., 2020). However, detailed insight into the nature and longevity of the Bmem cell compartment specific to SARS-CoV-2 is currently still unresolved (Vabret et al., 2020).

We extensively characterized the SARS-CoV-2-specific Bmem cell compartment using unique sets of fluorescently-labeled recombinant tetramers of the SARS-CoV-2 RBD and NCP antigens in combination with an extensive flowcytometry panel. The SARS-CoV-2-specific Bmem cells were quantified and characterized in 36 samples from 25 patients with COVID-19 or in convalescence. Circulating RBD- and NCP-specific Bmem cell subsets were detected early after infection and persisted over 242 days post-symptom onset. Early after infection, Ag-specific Bmem cells predominantly expressed IgM, over time followed by a predominance of IgG1. RBD-specific Bmem cell numbers were found to positively correlate with circulating Tfh cells suggesting prolonged germinal center (GC) activity. These analyses highlight that a decline in serum antibodies in convalescence may not reflect waning of immunity, but rather a contraction of the immune response with the development and persistence of B-cell memory.

## RESULTS

### Fluorescent NCP and RBD tetramers to identify SARS-CoV-2-specific B cells

The antigen-specific B-cell response to SARS-CoV-2 was characterized using recombinant forms of the RBD and NCP. Both proteins were generated in Expi293F cells with an AviTag for targeted biotinylation and tetramerization with fluorescently-labeled streptavidins to minimize epitope masking (**Figure 1A and B**). Two tetramers with distinct fluorochromes were generated for each protein: BV480 and BV650 for RBD, and BUV395 and BUV737 for NCP. In subjects with a history of COVID-19, distinct populations of RBD-specific and NCP-specific B cells were detected using double-discrimination (**Figure 1C**). Detection of these populations was highly specific, because neither population was detected in non-infected controls, and the RBD- and NCP-tetramers stained distinct B-cell subsets (**Figure 1C**).

**Figure 1.**
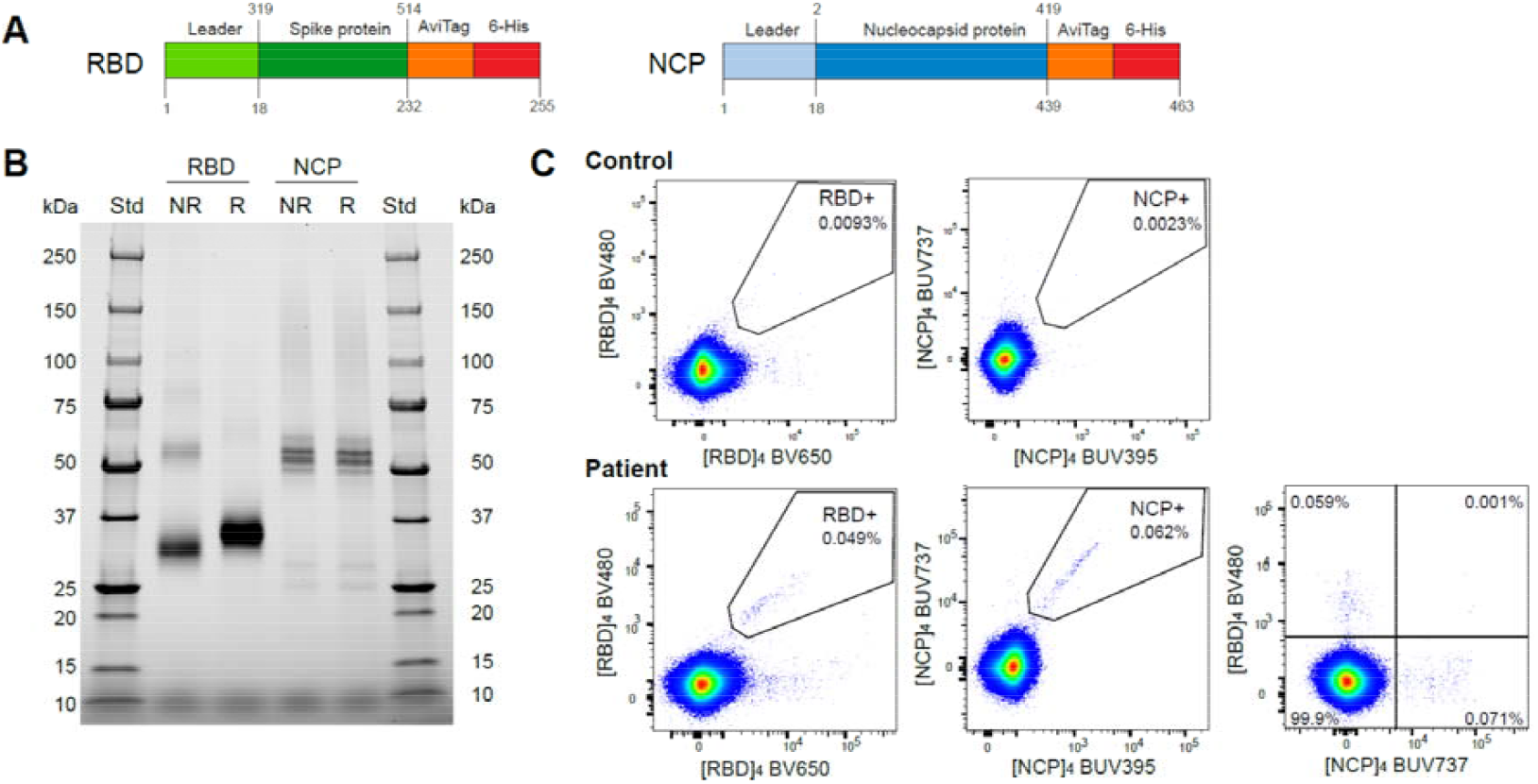
Construct design and detection of SARS-CoV-2 RBD- and NCP-specific B cells. **A)** Recombinant spike receptor binding domain (RBD) and nucleocapsid protein (NCP) constructs of SARS-CoV-2. **B)** SDS PAGE of purified, reduced (R) or non-reduced (NR) recombinant RBD and NCP. **C)** Flow cytometry stainings of CD19^+^ B cells from an uninfected control and a recovered COVID-19 patient using double discrimination through inclusion of two fluorescent tetramers for each protein (RBD or NCP) in the same staining tube. Percentages indicate the proportions of RBD- or NCP-specific cells within total CD19^+^ B cells.

### COVID-19 patient clinical and immunological characteristics

The SARS-CoV-2-specific antibody and B-cell response was investigated in 25 COVID-19 confirmed patients with samples obtained between 4- and 242-days after symptom onset (**Table 1**). Patients were classified into three levels of disease severity (WHO Working Group on the Clinical Characterisation and management of Covid-19 infection, 2020): six with severe disease requiring respiratory support in the intensive care unit (ICU); three with moderate disease requiring non-ICU hospital admission, and 16 with mild disease managed in the community (**Table 1**). 11 patients were sampled twice (paired samples); first between 21-106 days post-symptom onset and again at 116-242 days to evaluate the longevity of Bmem cell response to SARS-CoV-2 infection (**Table 1**). At the time of blood sampling, the majority of patients had normal blood counts of the major leukocyte and lymphocyte subsets (**Table 2**). Of note, the three patients sampled within 14 days post-symptom onset showed CD3^+^ T-cell lymphopenia due to reduced CD8^+^ T-cell counts (**Table 2**). All patients exhibited normal absolute numbers of B cells (**Table 2**).

**Table 1.** Patient characteristics.

| Patient | Age group (yrs) | Sex | Pre-existing conditions | Documented clinical features and symptoms | Treatment for COVID-19 | Disease classification* | Sampling time (Days post-symptom onset) |  |
| --- | --- | --- | --- | --- | --- | --- | --- | --- |
| 1 | 50-60 | F | none | Fever, dyspnoea, myalgia, fatigue, anosmia, ageusia | ICU, non-invasive ventilation, antivirals/immunomodulators | Severe | 4 |  |
| 2 | 60-70 | M | Diabetes mellitus | Dyspnea, cough, sore throat, rhinorrhea, fatigue, anosmia, loss of appetite | ICU, non-invasive ventilation, antivirals/immunomodulators | Severe | 12 |  |
| 3 | 60-70 | M | none | Fever, dyspnea, cough, myalgia, fatigue, confusion | ICU, non-invasive ventilation, antivirals/immunomodulators | Severe | 14 |  |
| 4 | 50-60 | M | Controlled asthma | Dyspnea, sedated (2 days post extubation) | ICU, invasive ventilation, antivirals/immunomodulators | Severe | 15 |  |
| 5 | 30-40 | M | none | Cough, headache, fatigue | Community managed | Mild | 21 | 119 |
| 6 | 20-30 | M | none | Cough, headache, arthralgias | Community managed | Mild | 22 | 116 |
| 7 | 20-30 | F | none | Fever, dyspnea, cough, headache, myalgias, anosmia, post-viral rash, tachycardia, congestion, fatigue | Community managed | Mild | 22 |  |
| 8 | 20-30 | F | none | Anosmia, ageusia, fatigue, nasal blockage | Community managed | Mild | 24 | 125 |
| 9 | 30-40 | M | none | Fever, cough, fatigue | Community managed | Mild | 26 | 120 |
| 10 | 30-40 | M | none | Cough, dyspnea, fever, sore throat, pneumonia, hypoxic respiratory failure | ICU, invasive ventilation, antivirals/immunomodulators | Severe | 30 |  |
| 11 | 30-40 | M | Type 2 diabetes mellitus, asthma, obesity | Fever, cough, dyspnea, anosmia, ageusia, myalgia, fatigue | Hospital general ward | Moderate | 43 | 129 |
| 12 | 30-40 | F | none | Asymptomatic | None | Mild | 43 | 129 |
| 13 | 20-30 | M | none | Cough, headache | Community managed | Mild | 50 |  |
| 14 | 20-30 | F | none | Dyspnea, headache, anosmia, fatigue | Community managed | Mild | 60 |  |
| 15 | 50-60 | M | none | Fever, cough, headache, myalgia, pneumonia, fatigue | Hospital general ward | Moderate | 69 | 163 |
| 16 | 30-40 | M | none | Fever, cough, headache | Community managed | Mild | 71 |  |
| 17 | 50-60 | F | SLE | Cough, sore throat, lower back pain | Community managed | Mild | 88 | 229 |
| 18 | 50-60 | M | none | Fever, cough, fatigue | Community managed | Mild | 92 | 228 |
| 19 | 20-30 | F | none | Influenza-like illness** | Community managed | Mild | 93 |  |
| 20 | 40-50 | F | none | Myalgia, fatigue | Community managed | Mild | 97 |  |
| 21 | 30-40 | M | none | Fever, dyspnea, headaches, myalgia, fatigue | Community managed | Mild | 98 | 232 |
| 22 | 30-40 | M | none | Fever, fatigue, anosmia, ageusia | Community managed | Mild | 106 | 242 |
| 23 | 60-70 | M | Ex-smoker, moderate alcohol intake | Fever, dyspnea, pneumonia, hypoxic respiratory failure | ICU, invasive ventilation and ECMO, antivirals/immunomodulators | Severe | 116 |  |
| 24 | 30-40 | M | none | Fever, sore throat, headache, tachycardia, hypoxia | Hospital general ward | Moderate | 132 |  |
| 25 | 40-50 | M | none | Dyspnea, cough, fatigue, chest tightness | Community managed | Mild | 186 |  |

\* Patients classified based on treatment for COVID-19: mild, community treated; moderate, hospital admission (non-intensive care); severe, hospital ICU and oxygen supplementation (WHO Working Group on the Clinical Characterisation and management of Covid-19 infection, 2020). \*\* No additional clinical details, symptoms and treatment details available. ECMO, Extracorporeal membrane oxygenation; SLE, systemic lupus erythematosus.

**Table 2.** Immunological details of patients.

| Patient | Days post-symptom onset | Disease classification | CD45<sup>+</sup> Leukocytes<br>x 10<sup>9</sup>/l | Granulocytes<br>cells/μl | Monocytes<br>cells/μl | Lymphocytes<br>cells/μl | B cells<br>cells/μl | NK cells<br>cells/μl | T cells<br>cells/μl | CD8<sup>+</sup> T cells<br>cells/μl | CD4<sup>+</sup> T cells<br>cells/μl |
| --- | --- | --- | --- | --- | --- | --- | --- | --- | --- | --- | --- |
| 1 | 4 | Severe | **2.5** | 1474 | **140** | **911** | 120 | 122 | **627** | **125** | 455 |
| 2 | 12 | Severe | 9.2 | 7089 | 721 | 1389 | 279 | 201 | **801** | 385 | 393 |
| 3 | 14 | Severe | <u>12.5</u> | <u>10915</u> | 742 | **797** | 152 | **42** | **555** | **130** | 371 |
| 4 | 15 | Severe | 6.6 | 4603 | 506 | 1464 | 331 | 51 | 1063 | 271 | 773 |
| 5 | 21 | Mild | 5.3 | 3249 | 398 | 1654 | 233 | 294 | 1060 | 281 | 727 |
|  | 119 |  | 4.5 | 2673 | 213 | 1611 | 224 | 53 | 1237 | 294 | 893 |
| 6 | 22 | Mild | **3.7** | 1684 | 300 | 1717 | 221 | 281 | 1245 | 438 | 744 |
|  | 116 |  | 5.2 | 2912 | 264 | 2060 | 377 | 128 | 1486 | 450 | 962 |
| 7 | 22 | Mild | **2.5** | **1168** | **124** | 1254 | 159 | 78 | 979 | 354 | 550 |
| 8 | 24 | Mild | 6.5 | 3783 | 377 | 2340 | 262 | 212 | 1767 | 536 | 1120 |
|  | 125 |  | 4.7 | 2313 | 180 | 2200 | 242 | 246 | 1570 | 471 | 972 |
| 9 | 26 | Mild | 4.9 | 2009 | 323 | 2568 | 315 | 210 | 2043 | 617 | 1172 |
|  | 120 |  | 6.1 | 2768 | 380 | 2987 | 461 | 139 | 2296 | 677 | 1383 |
| 10 | 30 | Severe | 5.4 | 2846 | 367 | 2187 | 181 | 309 | 1646 | 527 | 945 |
| 11 | 43 | Moderate | 6.3 | 3163 | <u>838</u> | 2300 | 286 | 369 | 1986 | <u>1209</u> | 732 |
|  | 129 |  | 6.5 | 3756 | 218 | 2511 | 205 | 251 | 1922 | <u>1612</u> | 690 |
| 12 | 43 | Mild | 6.1 | 3477 | 464 | 2159 | 171 | 179 | 1606 | 578 | 973 |
|  | 129 |  | 6.6 | 4177 | 207 | 2183 | 162 | 185 | 1778 | 672 | 1026 |
| 13 | 50 | Mild | 4.4 | 1650 | 576 | 2174 | 311 | 241 | 1839 | 630 | 1140 |
| 14 | 60 | Mild | 3.9 | 1837 | 182 | 1889 | 282 | 219 | 1227 | 327 | 824 |
| 15 | 69 | Moderate | 4.0 | 1824 | 364 | 1812 | 213 | 260 | 1340 | 669 | 634 |
|  | 163 |  | 6.1 | 3467 | 352 | 2244 | 402 | 267 | 1448 | 602 | 788 |
| 16 | 71 | Mild | 6.1 | 3056 | 439 | 2605 | 228 | 450 | 1632 | 652 | 881 |
| 17 | 88 | Mild | 4.1 | 2479 | 230 | 1402 | 193 | 163 | 1022 | 262 | 663 |
|  | 229 |  | 7.0 | 5203 | 238 | 1599 | 207 | 189 | 1176 | **237** | 755 |
| 18 | 92 | Mild | 4.3 | 2359 | 266 | 1640 | 198 | 254 | 1146 | 258 | 832 |
|  | 228 |  | **2.3** | **1006** | **59** | 1216 | 171 | 116 | 902 | 179 | 686 |
| 19 | 93 | Mild | 4.0 | 2148 | 218 | 1606 | 142 | 155 | 1276 | 406 | 816 |
| 20 | 97 | Mild | 4.0 | 2245 | 196 | 1586 | 134 | 274 | 1156 | 319 | 785 |
| 21 | 98 | Mild | **3.4** | 1796 | 308 | 1301 | 123 | 201 | 938 | **232** | 645 |
|  | 232 |  | **2.7** | **1396** | **152** | 1162 | 101 | 243 | **774** | **165** | 556 |
| 22 | 106 | Mild | **2.9** | **1321** | 226 | 1310 | 141 | 226 | 881 | 283 | 555 |
|  | 242 |  | **3.1** | **1445** | 210 | 1478 | 161 | 315 | **819** | **250** | 528 |
| 23 | 116 | Severe | 7.2 | 2227 | 313 | <u>4693</u> | 168 | <u>525</u> | <u>3835</u> | <u>1721</u> | <u>1719</u> |
| 24 | 132 | Moderate | 6.0 | 3500 | 611 | 1922 | 187 | 284 | 1370 | 632 | 637 |
| 25 | 186 |  | 3.8 | 2170 | 238 | 1388 | 168 | 151 | 1045 | 445 | 508 |
| Normal range: |  |  | **3.8-11.8** | **1454-8358** | **176-764** | **1140-3252** | **97-614** | **44-499** | **823-2430** | **257-973** | **348-1451** |

### COVID-19 patients generate neutralizing, RBD- and NCP-specific antibodies

The humoral response to SARS-CoV-2 infection in all patients was examined with a pseudovirus neutralization assay. 22/25 patients had neutralizing antibody titers to SARS-CoV-2, whereas none of the 36 uninfected controls had detectable neutralizing titers (**Figure 2A**). IgG ELISAs were performed to both SARS-CoV-2 RBD and NCP proteins in 25 patients and 36 controls. All 25 patients were positive for RBD-specific IgG and 24/25 were positive for NCP-specific IgG, i.e. 2 standard deviations (2SD) above the median of healthy controls (**Figure 2B and C**). All controls and patients had detectable levels of IgG to hemagglutinin (HA) of influenza H1N1 strain A/Michigan/45/2015 (**Figure 2D**) (Hartley et al., 2020), which was a recommended strain in the quadrivalent annual vaccine from 2017-2019 (Australian Immunisation Handbook, 2018). There was no significant difference in HA antibody levels between the patient and control groups. Thus, the recombinant SARS-CoV-2 RBD and NCP proteins are recognized by antibodies in COVID-19 patients with high sensitivity and specificity.

**Figure 2.**
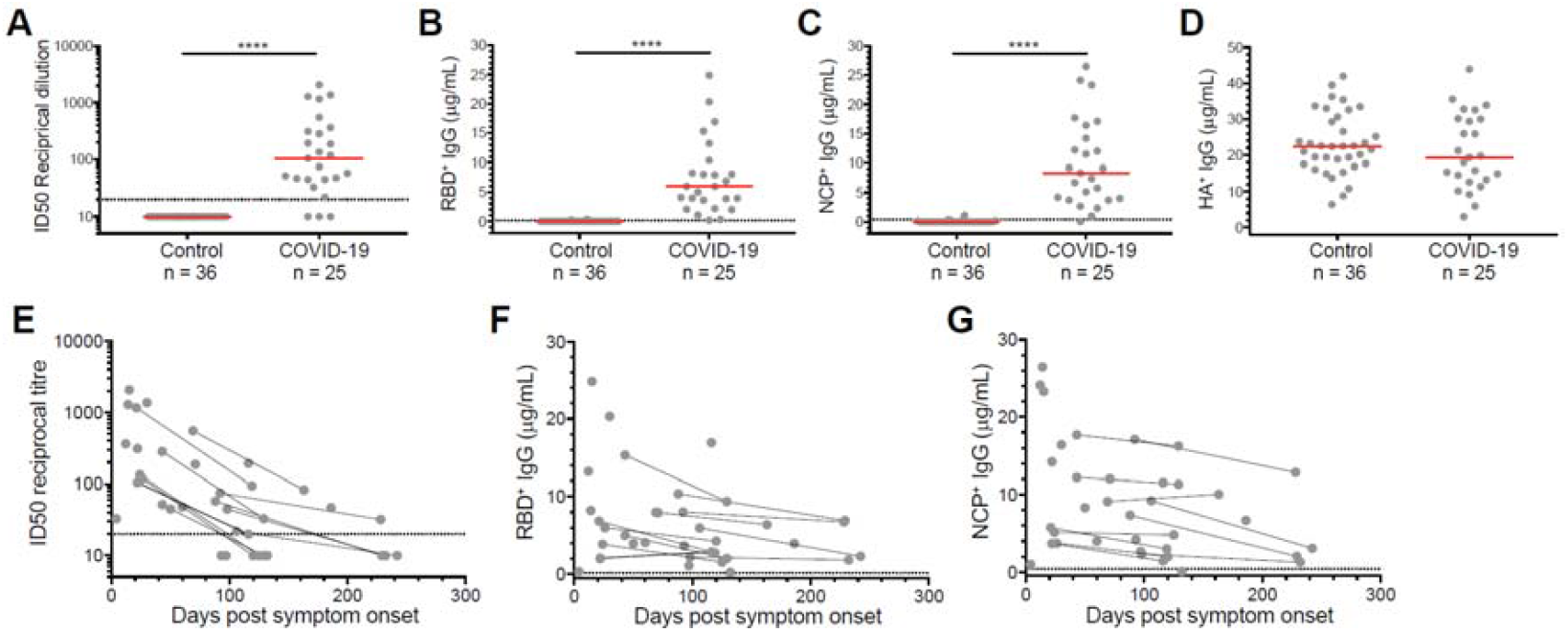
Neutralizing antibodies and RBD-, and NCP- and HA-specific IgG antibody levels. **A)** Neutralizing antibody titers to SARS-CoV-2 in 25 COVID-19 patients and 36 historic controls (sampled in 2019 and Q1 2020) as determined using a pseudovirus assay. Antigen-specific plasma IgG levels were determined to **B)** SARS-CoV-2 RBD, **C)** SARS-CoV-2 NCP and **D)** influenza A/Michigan/2015 haemagglutinin (HA). Horizontal solid red lines represent median values. **E)** Neutralizing antibody titers, and IgG levels to **F)** RBD and **G)** NCP plotted against time since symptom onset of infection of 25 patients including 11 patients sampled twice. The 11 paired samples are connected with black lines. The dotted horizontal lines in panels A and E depict an ID50 of 20, the cut-off for neutralization (Vietheer et al., 2017). The dotted horizontal lines in B, C, F and G depict the cut-off for positivity, defined as +2SD of the controls. Statistics were performed with the Mann-Whitney *U-*test for unpaired data; **** *p* ≤ 0.0001.

The neutralization titers (ID50), and RBD- and NCP-specific IgG levels in our patients declined over time in convalescence (**Figure 2E-G**). Neutralizing antibody titers were highest in patients sampled approximately 20 days post-symptom onset and subsequently contracted (**Figure 2E**). All ID50 titers were lower in the second sample of the 11 paired samples, and 7/11 repeat samples were at or below the threshold of neutralizing capacity (ID50 of 20) (**Figure 2E**). In parallel, RBD- and NCP-specific IgG levels were highest in the patients sampled around 20 days post-symptom onset, and in 10/11 repeat samples the RBD- and NCP-specific IgG levels were lower than the first draw (**Figure 2F and G**). Still, the decline after 20 days seemed to reach a plateau between 120-240 days with nearly all samples having detectable levels of RBD- and NCP-specific IgG.

### Detailed immune profiling of SARS-CoV-2-specific memory B cells

To examine the nature and kinetics of the RBD- and NCP-specific Bmem following SARS-CoV-2 infection, the RBD and NCP proteins were biotinylated and tetramerized with fluorescently-labeled streptavidins. RBD- and NCP-specific B cells were flowcytometrically evaluated in all 36 samples for expression of markers for plasmablasts (CD38), activated (CD71) and resting (CD27) Bmem cells, as well as surface IgD, IgA and IgG1, 2, 3 and 4 subclasses (**Figure 3A**) (**Supplementary Table 1**). Patients 1-3, sampled between 5-14 days post-onset of symptoms showed a large population of CD38^high^ CD27^+^ plasmablasts, whereas this population was negligible in any of the samples taken >20 days post-onset of symptoms (**Supplementary Figure 1**). Bmem cells were defined using IgD and CD27 (**Figure 3A-C**). All patients had detectable numbers of both IgG^+^ RBD- and NCP-specific Bmem cells, which were significantly higher than those of uninfected controls (*p* <0.0001 and *p* = 0.0005 respectively) (**Figure 3D**). The RBD- and NCP-specific Bmem cell populations contained both unswitched (CD27^+^IgM^+^IgD^+^) and Ig class-switched cells (CD27^+/-^IgD^-^) (**Figure 3B and C**). The latter subset predominantly contained IgG1-expressing Bmem cells with smaller proportions expressing IgG3 or IgA (**Figure 3E**). These distributions differed significantly between RBD- and NCP-specific Bmem cells: RBD-specific Bmem cells comprised significantly larger proportions of IgM^+^ IgD^+^, IgM only, IgG2 and total IgG expressing Bmem cell subsets than NCP-specific Bmem cells (**Figure 3E**). Compared to NCP-specific IgG^+^ Bmem cells, a higher proportion of RBD-specific IgG^+^ Bmem cells expressed CD27, a marker associated with increased replication and somatic hypermutation levels in Ig genes (**Figure 3F**) (Berkowska et al., 2011). Irrespective of the specificity, the proportions of IgG^+^ Bmem expressing CD27 were lower in patients sampled within 25 days post-symptom onset (**Supplementary Figure 2**). Thus, SARS-CoV-2 infection induces robust Bmem cell responses, which are predominantly comprised of IgM^+^ and IgG1^+^ Bmem cells with distinct immunophenotypes for those directed against RBD and NCP.

**Figure 3.**
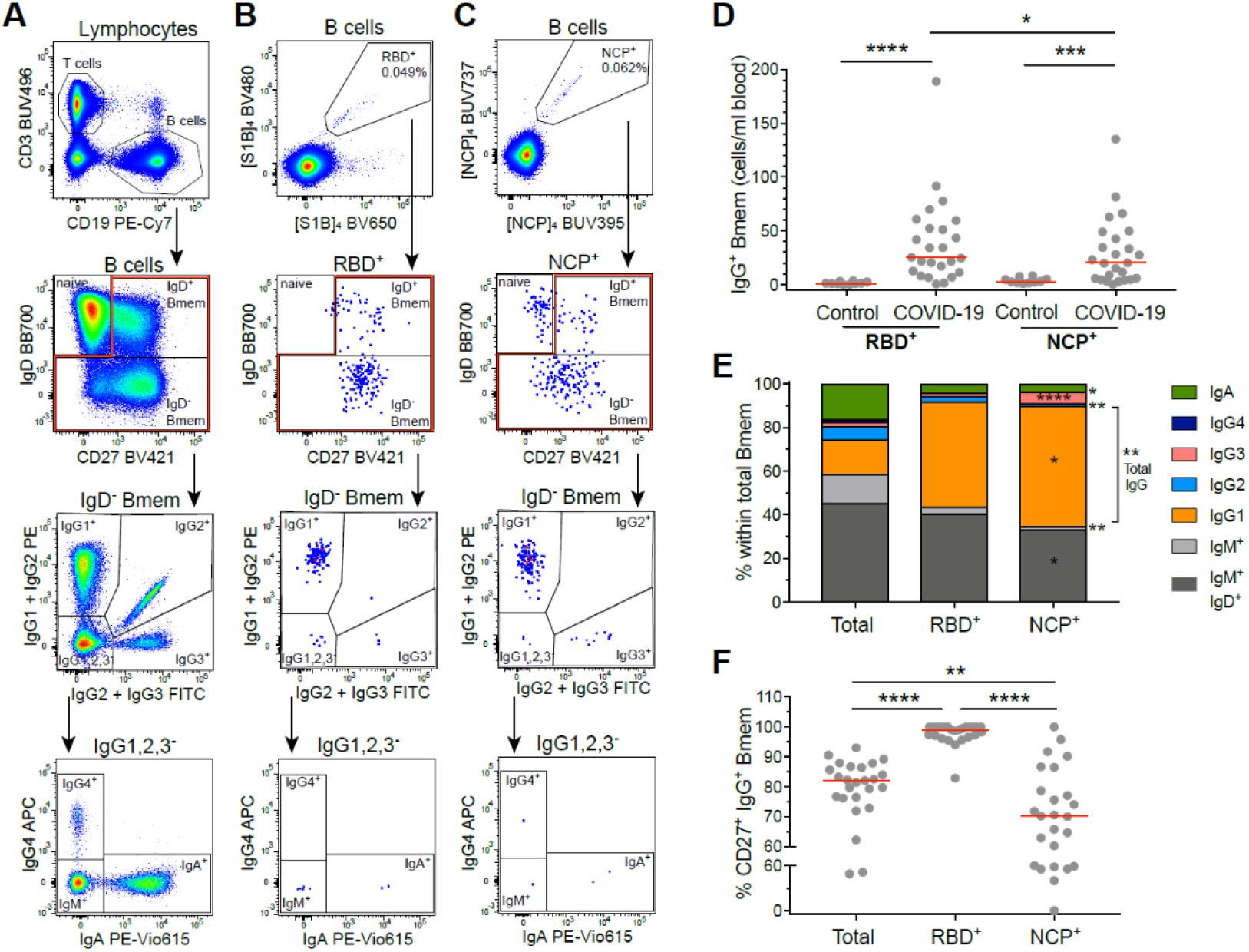
RBD- and NCP-specific Bmem cells predominantly express IgM or IgG1. **A)** Gating strategy to discriminate T and B cells, followed by subsetting of total B cells into CD27^-^IgD^+^ naive, CD27^+^IgD^+^ Bmem and CD27^+/-^IgD^-^ Bmem cells. Within IgD^-^ Bmem cells, Ig switched subsets were defined based on the differential expression of IgG1, 2, 3, 4 subclasses and IgA. **B)** Detection of RBD-specific (RBD^+^) B cells, and **C)** NCP-specific (NCP^+^) B cells, utilized the same gating strategy as for total B cells. **D)** Absolute numbers of IgG^+^ RBD^+^ and NCP^+^ Bmem cells in the first sample of 25 COVID-19 patients and 10 uninfected healthy controls. **E)** Median frequencies of total, RBD^+^ and NCP^+^ Bmem subsets in 25 COVID-19 patients. Significant differences between RBD^+^ and NCP^+^ Bmem subsets are depicted with asterisks in the NCP column. **F)** Frequencies of IgG^+^ Bmem cells expressing CD27 within total, RBD^+^ and NCP^+^ Bmem cells. Statistics: panel D, Mann-Whitney *U-*test for unpaired data; panels E and F, Wilcoxon matched-pairs signed rank test for paired samples; * *p* < 0.05, ** *p* < 0.01, *** *p* < 0.001, **** *p* < 0.0001.

### Long-term persistence of RBD- and NCP-specific Bmem expressing IgG

The numbers and Ig isotype distribution of RBD- and NCP-specific Bmem cell subsets varied between individuals. However, similar trends were still observed for both subsets with higher proportions and absolute numbers of IgG1^+^ RBD- and NCP-specific Bmem cells in samples taken 26 days or more post-symptom onset (**Figure 4A and B, Supplementary Figure 3**). RBD-specific Bmem cell numbers were highest between 100-150 days post-symptom onset (**Figure 4C**). Total and IgM^+^ Bmem cells in paired samples taken >200 days were lower than in the corresponding first samples, whereas IgG^+^ Bmem cells remained stable. NCP-specific Bmem cell numbers increased over the first 150 days as well, and in contrast to RBD-specific Bmem cells, they did not decline between 150-240 days (**Figure 4D**).

**Figure 4.**
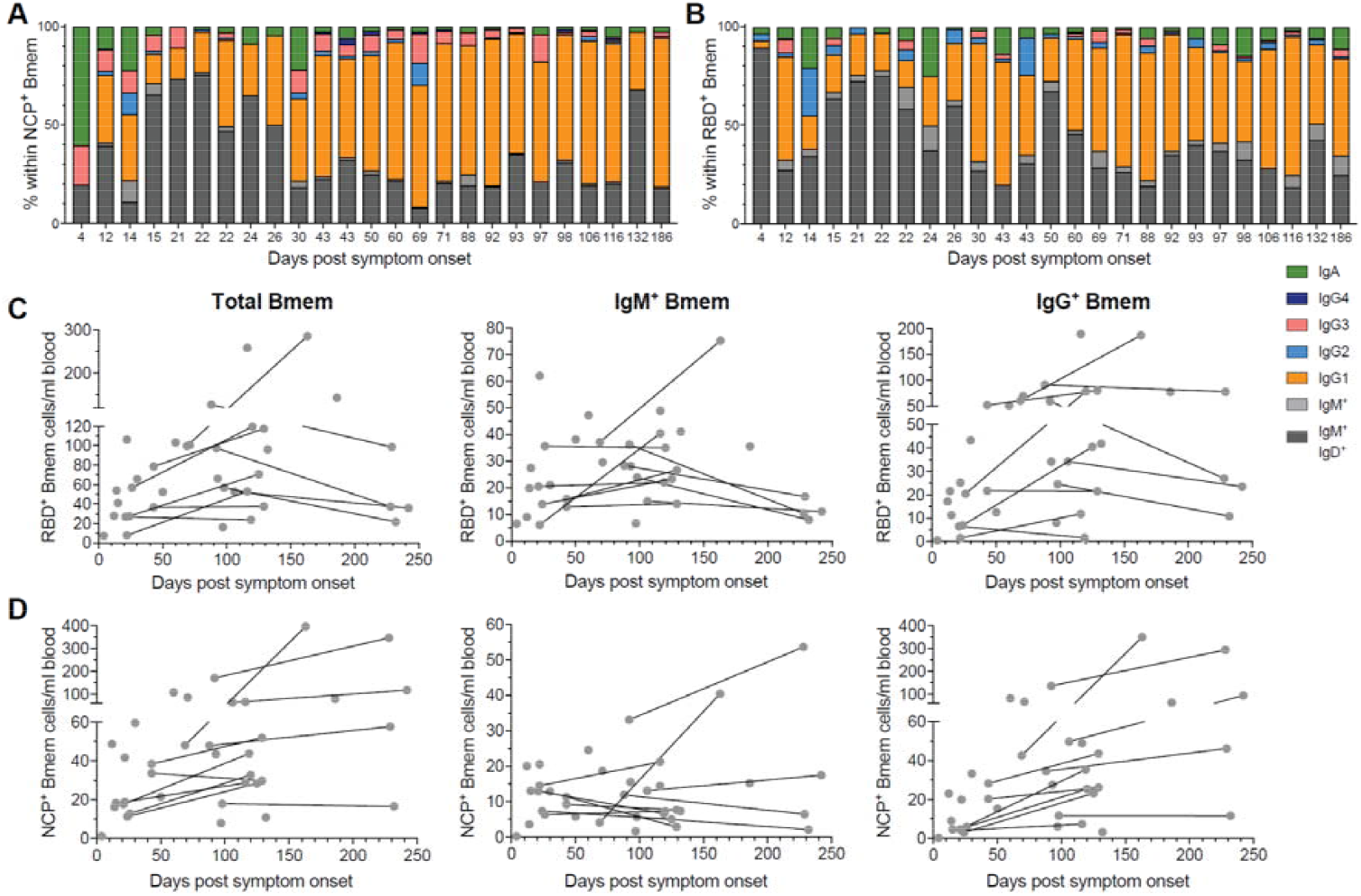
Composition and kinetics of SARS-CoV-2 RBD- and NCP-specific Bmem in convalescence. Relative composition of the Bmem cell compartment based on Ig isotype and IgG subclass expression subsets within **A)** RBD-specific (RBD^+^) and **B)** NCP-specific (NCP^+^) Bmem subsets. Patients’ data are ordered by days post-symptom onset. Absolute numbers of total, IgM^+^ and IgG^+^ Bmem cells specific for **C)** RBD^+^ or **D)** NCP^+^. Samples are plotted by days post-symptom onset for 25 individuals, with 11 patients sampled twice and paired samples connected with grey lines.

### Persistent germinal center activity in convalescent COVID-19 patients

IgM^+^ and IgG^+^ Bmem cells are predominantly generated in germinal center (GC) responses (Berkowska et al., 2011). To investigate for signs of ongoing GC activity beyond 50 days post-symptom onset, numbers of circulating follicular helper CD4^+^ T(fh)-cells, follicular regulatory CD4^+^ T(fr)-cells and CD8^+^ Tfh cells were examined. While total T-cell, CD4^+^ T-, CD8^+^ T- and γδ^+^ T-cell numbers did not change over time beyond 26 days post-symptom onset, the CD4^+^ Treg, CD4^+^ Tfh, CD4^+^ Tfr and the CD8^+^ Tfh subsets all trended to increase over the first 150 days followed by a plateau until 240 days (**Supplementary Figure 4**). Of these subsets, the CD4^+^ Tfh, CD4^+^ Tfr and CD8^+^ Tfh cell numbers showed a significant positive correlation with RBD-specific total, IgM^+^ and IgG^+^ Bmem cells across the 36 samples (**Figure 5A and B, Supplementary Figure 5**). In contrast, only CD8^+^ Tfh cell numbers showed a significant correlation with NCP-specific total and IgG^+^ Bmem cells (**Supplementary Figure 6**).

**Figure 5.**
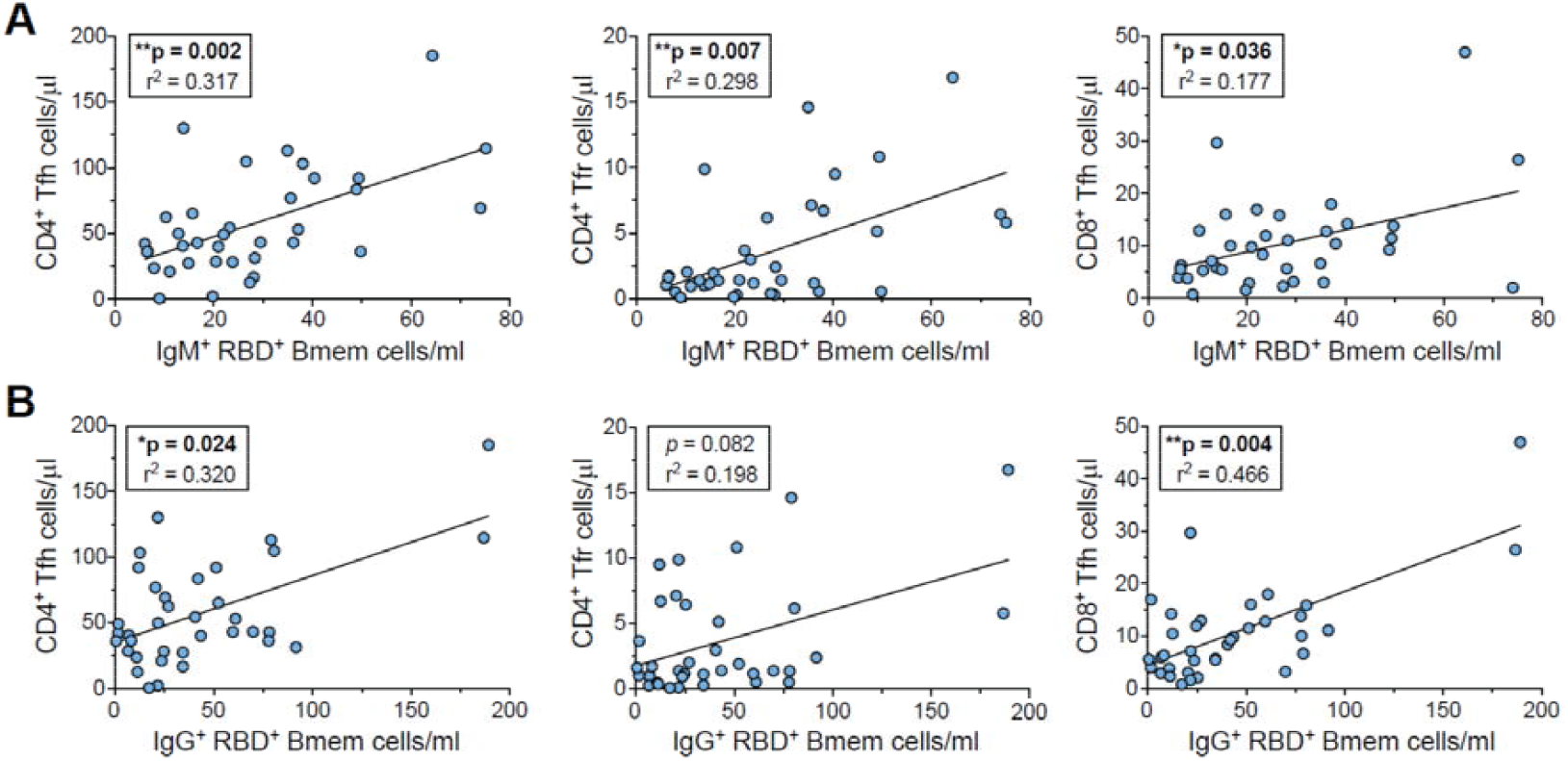
Positive correlations between RBD-specific Bmem numbers and CD4^+^ Tfh, CD4^+^ Tfr and CD8^+^ Tfh cells. **A)** RBD-specific IgM^+^ Bmem cell numbers plotted versus CD4^+^ Tfh, CD4^+^ Tfr and CD8^+^ Tfh cell numbers. **B)** RBD-specific IgG^+^ Bmem cell numbers plotted versus CD4^+^ Tfh, CD4^+^ Tfr and CD8^+^ Tfh cell numbers. Panels include all 36 samples from 25 patients. For population definitions, see **Supplementary Figure 4**. Trend lines depict linear correlations, statistics were performed using Spearman’s rank correlation; * *p* < 0.05, ** *p* < 0.01.

In summary, RBD- and NCP-specific IgG and Bmem cells were detected in all 25 patients with a history of COVID-19. While serum specific IgG levels declined with time post-symptom onset, SARS-CoV-2-specific B-cell numbers persisted, and RBD-specific Bmem cell numbers correlated with Tfh cell numbers. This long-term presence of circulating Bmem cell subsets directed against both the major SARS-CoV-2 neutralization target (RBD) and a non-neutralizing target (NCP) is indicative of long-term immunity following natural exposure and could provide a means to evaluate lasting immunity following vaccination.

## DISCUSSION

We have shown that COVID-19 patients rapidly generate B-cell memory to both the spike and nucleocapsid antigens following SARS-CoV-2 infection. Although rare events, numbers of Bmem cells specific for the major neutralization target, RBD, and a major non-neutralizing target, NCP, were both in the same order of magnitude. Using an extensive flow cytometry panel, we show that in line with typical T-cell dependent responses, IgM^+^ Bmem predominated in the first 20 days post-infection, followed by a gradual increase in IgG1^+^ Bmem. The fact that nearly all RBD-specific IgG^+^ Bmem cells expressed CD27 and their numbers correlated with circulating Tfh cells is indicative of long-lived immune memory.

In our patient population, we observed few abnormalities in the total B-cell population. All patients had total B-cell numbers within the normal range for age-matched controls. This finding is in contrast to some studies that reported a decrease in B-cell frequencies during COVID-19 infection which normalized in convalescence (Kaneko et al., 2020; Mathew et al., 2020; Oliviero et al., 2020; Zhou et al., 2020). However, these studies report frequencies rather than absolute numbers and as such an increase in other immune subsets such as NK cells could cause the observed decrease in B- and T-cells (Mathew et al., 2020; Zhou et al., 2020).

Patients sampled within 14 days of symptom onset had an enlarged plasmablast population that was absent in those sampled beyond 20 days post-symptom onset and who are in convalescence. Others have identified this transient plasmablast expansion in COVID-19 (Mathew et al., 2020; Newell et al., 2020). However, this observation is not restricted to SARS-CoV-2 infection and has been reported in other viral infections including influenza and dengue virus (Huang et al., 2014; Wrammert et al., 2012). Quantification of total B-cell subsets is important for the monitoring of disease progression and reconstitution of the humoral response post-infection. However, whilst alterations seen in SARS-CoV-2 infection at this level can be indicative of severity of infection or clinical disease, they are not unique and hence not indicative of long-lived SARS-CoV-2-specific humoral responses.

The detection of virus-specific antibodies is regularly employed in the diagnosis of other viral infections including Epstein-Barr virus, Cytomegalovirus, Hepatitis B virus, and Influenza virus (Gulley and Tang, 2008; Krajden et al., 2005; Nhat et al., 2017; Ross et al., 2011). The antibody response to SARS-CoV-2 infection has been shown to be directed to multiple targets of the virus including the spike, with those that target the RBD considered neutralizing (Gudbjartsson et al., 2020; Long et al., 2020; Ng et al., 2020; Vabret et al., 2020). Other antibodies target the NCP (Gudbjartsson et al., 2020; Long et al., 2020; Ng et al., 2020) or non-structural proteins (Hachim et al., 2020). Detection of such antibodies can be used as markers of recent infection. However, it has been reported that antibody levels to SARS-CoV-2 decrease over time (Gudbjartsson et al., 2020; Huang et al., 2020; Ng et al., 2020). What we now show is that this decrease reflects a contraction of the immune response. Despite the decline in antibody levels, these remain present at detectable levels until 240 days post-symptom onset and their presence is accompanied by the persistence of RBD- and NCP-specific Bmem cells. These antigen-specific Bmem are rapidly generated and particularly those expressing IgG^+^ remain numerically high, and hence may represent a more robust marker of long-term immunity.

Antigen-specific Bmem cells are very rare events at 0.008-0.1% of B cells. Here, we used double-discrimination to exclude B cells that non-specifically bound to the fluorochrome, which is typically observed when utilizing large protein-based fluorochromes (e.g. phycoerythrin (PE) and allophycocyanin (APC)) (Brouwer et al., 2020; Juno et al., 2020; Kaneko et al., 2020; Liu et al., 2019; Rodda et al., 2020; Wheatley et al., 2016; Zhou et al., 2020). To further overcome this limitation, we used non-protein polymer fluorochromes, which exhibited minimal non-specific B-cell binding and increased the sensitivity of our assay (Hartley et al., 2020).

Our study, for the first time shows the kinetics and longevity of SARS-CoV-2-specific Bmem cell numbers. Other studies have identified SARS-CoV-2-specific B cells in COVID-19 patients with particular focus on the RBD of the spike protein, mainly for the purpose of cloning neutralizing SARS-CoV-2-specific antibodies (Juno et al., 2020; Nguyen-Contant et al., 2020; Robbiani et al., 2020; Rogers et al., 2020; Wilson et al., 2020). Such studies have observed a predominant IgG^+^ B-cell response to SARS-CoV-2 with lower frequencies of cells expressing IgM and IgA (Brouwer et al., 2020; Juno et al., 2020; Rodda et al., 2020). We have expanded on this through detailed flow cytometry with the inclusion of absolute cell counts to show that SARS-CoV-2-specific Bmem cells predominantly expressed IgM or IgG1. This distinction enabled the discrimination between the initially large fraction of IgM^+^ Bmem cells that tended to decline beyond 150 days, while the IgG expressing fraction persisted and those specific for RBD correlated strongly with circulating Tfh cells.

The Bmem cell populations directed against the two SARS-CoV-2 targets showed remarkable differences. The RBD-specific Bmem cells were nearly all CD27^+^ and strongly correlated with Tfh cells, while this was not observed for NCP-specific Bmem cells. Differential expression of CD27 and IgG subclasses on human Bmem cells are associated with different maturation stages (Berkowska et al., 2011; de Jong et al., 2017). Typically, CD27^+^ Bmem cells have more somatic hypermutations and have undergone more cell divisions than those lacking CD27 expression (Berkowska et al., 2011; de Jong et al., 2017). Thus, the increased proportions of RBD-specific IgG^+^ Bmem cells expressing CD27 as compared to NCP-specific IgG^+^ Bmem cells could suggest that that RBD-specific Bmem cells may remain longer in the GC. This is further supported by the fact that only the RBD-specific Bmem cell numbers correlated with CD4^+^ and CD8^+^ Tfh cells. Limited GC activity has been reported in COVID-19 patients early in convalescence (15-36 days post-infection) (Kaneko et al., 2020). It would be of interest to study whether this activity increases or persists beyond 36 days. Similarly, detailed molecular studies and somatic hypermutation analysis of RBD-specific Bmem could provide insights into prolonged GC activity. For ongoing GC reactions, continual antigen exposure is required. Human coronaviruses, including SARS-CoV-2, form double membrane vesicles in the host cell as part of their replication cycle (Angelini et al., 2013; Snijder et al., 2020). These vesicles contain antigen (Snijder et al., 2006), persist after infection and hence, could drive prolonged GC activity.

We here show long-term persistence of SARS-CoV-2-specific Bmem cells with kinetics that suggest high durability, particularly of those expressing IgG. Bmem cells show evidence of antigen experience through extensive replication cycles, elevated somatic hypermutation levels and class switch recombination (Berkowska et al., 2011; de Jong et al., 2017). After infection, a portion of Bmem cells can be measured with an ‘activated’ phenotype (CD21^lo^ CD27^+^) and this population has been shown to contract after ∼2 weeks (Andrews et al., 2019; Ellebedy et al., 2016). The remaining Bmem cells are defined as ‘resting’ with a predominant CD21^+^ CD27^+^ phenotype. Furthermore, Bmem cells have been shown to increase expression of surface molecules including CD80, CD180 and TACI indicating the potential for rapid activation upon antigen re-encounter (Berkowska et al., 2011; Berkowska et al., 2015). Whilst these markers have not been assessed in SARS-CoV-2-specific Bmem cells, they display the classical surface markers (ie. CD27^+/-^ IgG^+^) indicating long-lived memory. There have been some studies reporting that the Bmem cell response to SARS-CoV may not be long-lived (Tang et al., 2011; Traggiai et al., 2004), however, our results indicate that SARS-CoV-2 infection generates long-lasting B-cell immunity up to 8 months post-infection and could be protective upon reinfection.

In this study, we sampled peripheral blood and hence measured the systemic Bmem cell response to SARS-CoV-2 infection. We know from vaccination studies in mice and humans that local and systemic Bmem cells are phenotypically different (Allie et al., 2019; Koutsakos et al., 2018). It has also been shown that influenza-specific Bmem cells persist in the lungs of mice and do play a role in protection upon reinfection (Adachi et al., 2015; Onodera et al., 2012). However, at present the attributes of SARS-CoV-2 immunity in the respiratory tract are largely unknown. As knowledge of SARS-CoV-2 and human lung immunology evolve, we will gain insight into what is required for a protective response to this respiratory virus. However, we propose that the establishment of systemic immunity will prevent severe systemic COVID-19, and reinfection may be limited to a mild or asymptomatic upper respiratory tract infection.

The identification and analysis of SARS-CoV-2-specific Bmem cells could potentially be used as a surrogate marker of humoral immunity in vaccination studies. Currently, SARS-CoV-2 vaccination trials investigate predominantly SARS-CoV-2-specific and neutralizing antibodies as markers of vaccine efficacy (Corbett et al., 2020; Folegatti et al., 2020; Gao et al., 2020; Logunov et al., 2020; Mercado et al., 2020; Wang et al., 2020). As we have shown that SARS-CoV-2-specific Bmem cell numbers are stable over time, we propose that they may represent a more robust marker of long-lived humoral immunity compared to serum antibodies. As antibody levels decline when the immune response contracts, IgG^+^ Bmem cells remain present, indicating that SARS-CoV-2 induces long-term humoral immunity. Therefore, cellular measurements of immune response could be more reliable markers for immunity following natural infection or vaccination.

## METHODS

### Participants

Individuals with a PCR-confirmed diagnosis of COVID-19 and uninfected controls were enrolled in research studies to examine their peripheral blood B- and T-cell subsets (projects: Alfred Health Human Research and Ethics Committee Numbers 182/20 and 202/20, Monash University 2016-0289 and 2020-26385). From March to September 2020, 25 individuals with a history of PCR-confirmed COVID-19 disease and 36 healthy controls (sampled in 2019 and Q1 2020) consented to one or two 40 ml donations of blood as well as the collection of clinical data including: basic demographics (age, sex), clinical details of COVID-19 (clinical symptoms, date of symptom onset, and COVID-19 specific treatments) and co-morbid medical conditions (**Table 1**). This study was conducted according to the principles of the Declaration of Helsinki and approved by local human research ethics committees.

### Sample processing

Blood samples of patients and controls were processed as previously described (Edwards et al., 2019; Hartley et al., 2020). Briefly, 200 µl was used for whole blood cell counts (Cell Dyn analyser; Abbott core laboratory, Abbott Park, IL) and Trucount analysis (see flow cytometry section). The remainder was used to separate and store plasma (−80°C), and to isolate live peripheral blood mononuclear cells (PBMC) by Ficoll-paque density gradient centrifugation and cryopreservation at a cell density of 10 million cells/ml in RPMI medium with 40% FCS and 10% DMSO in liquid nitrogen for later analysis of SARS-CoV-2-specific B cells.

### Protein production and tetramerization

Recombinant nucleocapsid protein (NCP) and receptor binding domain of the spike protein (RBD) of SARS-CoV-2 were produced with a human immunoglobulin (Ig) leader and the Fel d 1 leader sequence, respectively. Each protein construct was C-terminally fused to the biotin ligase (BirA) AviTag target sequence and a 6His affinity tag (**Figure 1A**). The DNA constructs were cloned into a pCR3 plasmid and produced and purified as described previously (Hartley et al., 2020). Briefly, plasmid DNA was purified from *E. coli* by Maxiprep (Zymo Research, Irvine, CA), and 30 μg DNA was transfected into 293F cells using the Expi293 Expression system (Thermo Fisher, Waltham, MA). Supernatants from 25 ml cultures were collected on days 3 (NCP) or 5 (RBD) post-transfection and purified by application to a Talon NTA-cobalt affinity column (Takara Bio, Kusatsu, Shiga, Japan) with elution in 200 mM Imidazole. Eluted proteins were then dialyzed against 10 mM Tris for 48 hours at 4°C. Purified proteins were biotinylated by incubating at room temperature overnight with 1/8 of final volume each of Biomix A (0.5M Bicine-HCl, pH8.3) and Biomix B (100mM ATP, 100mM MgOAc, 500 μM D-biotin) followed by 2.5 μg of BirA enzyme per milligram of protein. Biotinylated protein was subsequently dialyzed against 10 mM Tris for 36 hours at 4°C, and subsequently stored at -80°C prior to use. Soluble biotinylated NCP protein was tetramerized by the addition of either Brilliant Ultra Violet (BUV)395-conjugated streptavidin, or streptavidin-BUV737, and biotinylated RBD with streptavidin-BV480 or streptavidin-BV650 (BD Biosciences, Franklin lakes, NJ) at a protein:streptavidin molar ratio of 4:1 making 4 unique tetramers: [NCP]_4_-BUV395, [NCP]_4_-BUV737, [RBD]_4_-BV480 and [RBD]_4_-BV650.

### SDS-PAGE

SDS-PAGE analyses were performed as described previously (Hartley et al., 2020). Briefly, 10 μl of sample was mixed with 2.5 μl of 4X Laemil Sample buffer (non-reducing) (BioRad, Hercules, CA) or reducing buffer (4X Laemil Sample buffer with the addition of 1.25 μl DTT). Samples under reducing conditions were heated to 85°C for 10 minutes. 10 μl of ladder (1:1 mixture) of Precision plus protein standard (Unstained and All blue, both from BioRad), and reduced or non-reduced sample was loaded on a 4-15% Mini-PROTEAN TGX Stain-Free gel (BioRad) and run for 30 minutes at 200 V then imaged on the BioRad ChemiDoc Touch imaging system (BioRad).

### Measurement of SARS-CoV-2 neutralizing antibodies in plasma

Measurement of neutralizing antibodies was performed using SARS-CoV-2 retroviral pseudotyped particles and a 293T-ACE2 cell line (Crawford et al., 2020) as described before (Jackson et al., 2020). Plasma was heat inactivated at 56°C for 45 minutes followed by serial dilution in DMF10. Each dilution was mixed in duplicate with an equal volume of SARS-CoV-2 (WUHAN-1 spike) retroviral pseudotyped virus and incubated for 1 hour at 37°C. Virus-plasma mixtures were added to 293T-ACE2 cell monolayers in 96-well poly-L-lysine coated plates seeded the day prior at 10,000 cells/well, and incubated for 2 hours at 37°C before addition of an equal volume of DMF10 and incubated. After 3 days, tissue culture fluid was removed, monolayers were washed once with PBS and lysed with cell culture lysis reagent (Promega, Madison, WI) and luciferase measured using luciferase substrate (Promega) in a Clariostar plate reader (BMG LabTechnologies, Offenburg, Germany). The mean percentage entry was calculated as (relative light units (RLU) plasma+virus)/(RLU medium+virus)*100. The percentage entry was plotted against the reciprocal dilution of plasma in GraphPad Prism 8 Software (GraphPad Software, La Jolla, CA) and curves fitted with a one-site specific binding Hill plot. The reciprocal dilution of plasma required to prevent 50% virus entry was calculated from the non-linear regression line (ID50). The lowest amount of neutralizing antibody detectable is a titer of 20 (Vietheer et al., 2017). All samples that did not reach 50% neutralization were assigned an arbitrary value of 10.

### ELISA

EIA/RIA plates (Costar, St Louis, MO) were coated with 2 μg/ml recombinant SARS-CoV-2 NCP or RBD or with hemagglutinin (HA) from influenza A/Michigan/08/2015 (AM15) overnight at 4°C (Hartley et al., 2020). Plates were subsequently blocked with 3% BSA in PBS and incubated with diluted plasma samples, 1:30 for RBD and NCP and 1:50 for AM15. Antigen-specific IgG was detected by adding rabbit anti-human IgG HRP (Dako, Glostrup, Denmark). ELISA plates were developed using TMB solution (Life Technologies, Carlsbad, CA) and the reaction was stopped with 1 M HCl. Absorbance (OD450nm) was measured using a Multiskan Microplate Spectrophotometer (Thermo Fisher). Serial dilutions of recombinant human IgG (in-house made human Rituximab) in separate wells on the same plate were performed for quantification of specific IgG.

### Flow cytometry

Absolute numbers of leukocyte subsets were determined as previously described (Edwards et al., 2019; Hartley et al., 2020). Briefly, 50 μl of whole blood was added to a Trucount tube (BD Biosciences) together with a 20 μl antibody cocktail containing antibodies to CD3, CD4, CD8, CD16, CD19, CD56 and CD45 (**Supplementary Tables 1 and 2**) and incubated for 15 minutes at room temperature in the dark. Subsequently samples were incubated for a further 15 minutes at room temperature with 500 μl of 0.155 M NH_4_Cl to lyse red blood cells. The tube was then stored in the dark at 4°C for up to 2 hours prior to acquisition on the LSRII analyzer (BD Biosciences).

Detailed T-cell subsetting was performed with two 11-color flow cytometry panels (**Supplementary Table 1**) as previously described (Edwards et al., 2019). Specific subsets were defined as follows: CD4^+^ Treg, CD3^+^CD4^+^CD8^-^CD127^-^CD25^+^; CD4^+^ Tfh, CD3^+^CD4^+^CD8^-^CD45RA^-^CXCR5^+^; CD4^+^ Tfr, CD3^+^CD4^+^CD8^-^CD127^-^CD25^+^ CD45RA^-^CXCR5^+^; CD8^+^ Tfh, CD3^+^CD4^-^CD8^+^CD45RA^-^CXCR5^+^ (**Supplementary Figure 4**).

For detection of antigen-specific B cells, 12.5 million PBMC were incubated with fixable viability stain 700 (BD Biosciences), antibodies against CD3, CD19, CD21, CD27, CD38, CD71, IgA, IgD, IgG1, IgG2, IgG3, IgG4, (**Supplementary Tables 1 and 2**) and 5 μg/ml (total of 1.25 ug per 250 µl stain) each of [NCP]_4_-BUV395, [NCP]_4_-BUV737, [RBD]_4_-BV480 and [RBD]_4_-BV650 for 15 minutes at room temperature in a total volume of 250 μl FACS buffer (0.1% sodium azide, 0.2% BSA in PBS). In addition, 5 million PBMC were similarly incubated with fixable viability stain 700 (BD Biosciences), antibodies against CD3, CD19, CD27 and IgD, plus BUV395-, BUV737-, BV480- and BV650-conjugated streptavidin controls (**Supplementary Tables 1 and 2**). Following staining, cells were washed twice with FACS buffer and filtered through a 70 μM filter prior to acquisition on the 5-laser BD LSRFortessa X-20. Flow cytometer set-up and calibration was performed using standardized EuroFlow SOPs, as previously described (**Supplementary Tables 3 and 4**) (Edwards et al., 2019; Hartley et al., 2020; Kalina et al., 2012).

### Data analysis and statistics

All flow cytometry data was analyzed with FlowJo v10 software (TreeStar, Ashland, Ore). Statistical analysis was performed with GraphPad Prism 8 Software (GraphPad Software). Matched pairs were analyzed with the non-parametric Wilcoxon matched pairs signed rank test. Unpaired groups were analyzed with the non-parametric Mann-Whitney *U*-test. Correlations were performed using the non-parametric Spearman’s rank correlation. For all tests, *p* < 0.05 was considered significant.

## Data Availability

All research data are included and available in the Tables, Figures and supplemental data of this manuscript

## ACKNOWLEDGEMENTS

We gratefully acknowledge Dr. Bruce D. Wines, Ms. Sandra Esparon for technical assistance, Ms Kirsten Deckert, Ms Jessica Wisniewski and Ms Anna Coldham for their support with blood patient sampling and E/Prof Jennifer Rolland for critical evaluation of the manuscript. The authors thank Marsha Hartman from BD Biosciences for unrestricted in-kind provision of reagents early in the pandemic to create a solution for standardized approaches to vaccine development and clinical research. AYP is supported by an Australian National Health and Medical Research Council Practitioner Fellowship. MCvZ is supported by a Senior Research Fellowship (1117687) from the Australian National Health and Medical Research Council.

## AUTHOR CONTRIBUTIONS

GEH designed and performed experiments, analyzed the data, and wrote the manuscript. ESJE designed experiments, analyzed the data, supervised the work and edited the manuscript. PMA and NV performed experiments and analyzed the data. SJ recruited patients and extracted clinical information. JM and AYP established the Alfred Biobank and edited the manuscript. IB conducted neutralization assays. HED conducted neutralization assays and edited the manuscript. PMH designed experiments and edited the manuscript. ROH designed experiments, supervised the work and edited the manuscript. MCvZ designed experiments, analyzed the data, supervised the work, and wrote the manuscript.

