## Supplemental data for "Rapid and lasting generation of B-cell memory to SARS-CoV-2 spike and nucleocapsid proteins in COVID-19 disease and convalescence"

**Supplementary Tables (n= 4) and Figures (n=6)**

**Supplementary Table 1. Composition of the antibody panels**

|  | **Fluorochrome** | | | | | | | | | | | | | | | | |
| --- | --- | --- | --- | --- | --- | --- | --- | --- | --- | --- | --- | --- | --- | --- | --- | --- | --- |
| Tube | **BUV395** | **BUV496** | **BUV737** | **BV421** | **BV480/**  **BV510** | **BV605** | **BV650** | **BV711** | **BV786** | **FITC/ BB515** | **PerCP-Cy5.5/ BB700** | **PE** | **PE- CF594/ PE-Vio615** | **PC7/**  **PE-Cy7** | **APC** | **AF700** | **APC-Cy7 / APC-H7** |
| 1. TruCount | **-** | **-** | **-** | **-** | **-** | **-** |  | **-** | - | CD3 | CD45 | CD16 + CD56 | **-** | CD4 | CD19 | - | CD8A |
| 2. Ag-specific Bmem | NCP | CD3 | NCP | CD27 | RBD | - | RBD | CD21 | CD71 | IgG2 + IgG3 | IgD | IgG1 + IgG2 | IgA | CD19 | IgG4 | Viability | CD38 |
| 3. Streptavidin control | Strep | - | Strep | CD27 | Strep | - | Strep | - | - | CD3 | IgD | - | - | CD19 | - | Viability | - |
| 4. T-effector | - | - | - | CD27 | CD4 | CD45RA | - | CD3 | - | CD45RO | CD28 | CD31 | CCR7 | TCRγδ | HLA-DR | - | CD8A |
| 5. Th subset | - | - | - | CD25 | CD4 | CD45RA | - | CD3 | - | CXCR5 | CCR6 | CXCR3 | CCR7 | CCR4 | CD127 | - | CD8A |

**Supplementary Table 2. Antibody list**

| **Marker** | **Fluorochrome** | **Clone** | **Source** | **Cat. number** | | **Volume/**  **test (μl)** | **Tube(s)** |
| --- | --- | --- | --- | --- | --- | --- | --- |
| CD3 | BUV496 | UCHT1 | BD Bioscience | 612940 | 1 | | 2 |
| CD3 | FITC | UCHT1 | BD Biosciences | 555332 | 3 / 1 | | 1 / 3 |
| CD3 | BV711 | UCHT1 | BD Biosciences | 563725 | 2.5 | | 4 / 5 |
| CD4 | PC7 | SFCI12T4D11 | Beckman Coulter | 6607101 | 0.2 | | 1 |
| CD4 | BV510 | RPA-T4 | Biolegend | 300546 | 1.5 | | 4 / 5 |
| CD8A | APC-H7 | SK1 | BD Biosciences | 560179 | 4 | | 1 |
| CD16 | PE | B73.1 | Biolegend | 360704 | 0.2 | | 1 |
| CD19 | APC | SJ25C1 | Biolegend | 363006 | 0.4 | | 1 |
| CD19 | PE-CY7 | SJ25C1 | BD Biosciences | 557835 | 5 | | 2 |
| CD21 | BV711 | B-ly4 | BD Biosciences | 563163 | 5 | | 2 |
| CD25 | BV421 | BC96 | Biolegend | 302630 | 2.5 | | 5 |
| CD27 | BV421 | M-T271 | BD Biosciences | 562513 | 1 | | 2 / 3 |
| CD28 | PerCP-Cy5.5 | CD28.8 | Biolegend | 302922 | 5 | | 4 |
| CD31 | PE | WM59 | BD Biosciences | 555446 | 5 | | 4 |
| CD38 | APC-Cy7 | HIT2 | Biolegend | 303534 | 0.2 | | 2 |
| CD45 | PerCP-Cy5.5 | 2D1 | BD Biosciences | 340953 | 2 | | 1 |
| CD45RA | BV605 | HI100 | Biolegend | 304134 | 0.2 | | 4 / 5 |
| CD45RO | FITC | UCHL1 | Biolegend | 304204 | 5 | | 4 |
| CD56 | PE | B159 | BD Biosciences | 555516 | 5 | | 1 |
| CD71 | BV786 | M-A712 | BD Biosciences | 563768 | 1 | | 2 |
| CD127 | APC | A019D5 | Biolegend | 351316 | 5 | | 5 |
| CCR4 | PE-CY7 | L291H4 | Biolegend | 359410 | 5 | | 5 |
| CCR6 | PerCP-CY5.5 | G034E3 | Biolegend | 353406 | 2.5 | | 5 |
| CCR7 | PE CF594 | 150503 | BD Biosciences | 562381 | 5 | | 4 / 5 |
| CXCR3 | PE | 1C6/CXCR3 | BD Biosciences | 557185 | 20 | | 5 |
| CXCR5 | BB515 | RF8B2 | BD Biosciences | 564624 | 5 | | 5 |
| HLA-DR | APC | L243 | Biolegend | 307610 | 2 | | 4 |
| IgA | PE-Vio615 | REA1014 | Miltenyi Biotec | 130-116-882 | 1.5 | | 2 |
| IgD | BB700 | IA6-2 | BD Biosciences | 566538 | 1 | | 2 |
| IgG1 | PE | SAG1 | Cytognos | CYT-IGG1PE | 1 | | 2 |
| IgG2 | FITC | SAG2 | Cytognos | CYT-IGG2F | 2 | | 2 |
| IgG2 | PE | SAG2 | Cytognos | CYT-IGG2PE | 2 | | 2 |
| IgG3 | FITC | SAG3 | Cytognos | CYT-IGG3F | 2 | | 2 |
| IgG4 | APC | SAG4 | Cytognos | CYT-IGG4AP | 2 | | 2 |
| IgM | BV510 | MHM-88 | Biolegend | 314522 | 1 | | 2 / 3 |
| Strep | BUV395 | - | BD Biosciences | 564176 | 0.36 | | 3 |
| Strep | BUV737 | - | BD Biosciences | 564293 | 0.36 | | 3 |
| Strep | BV480 | - | BD Biosciences | 564876 | 0.67 | | 3 |
| Strep | BV650 | - | Biolegend | 405232 | 0.13 | | 3 |
| TCRγδ | PC7 | IMMU510 | Beckman Coulter | B10247 | 1 | | 4 |
| Viability | AF700 | - | BD Biosciences | 564997 | 0.1 | | 2 / 3 |

**Supplementary Table 3. Flow cytometer set-up**

| **LSRFortessa X-20** | | **LSRII** | | **Fluorochromes used in this study** |
| --- | --- | --- | --- | --- |
| **355 nm** | | **-** | |  |
| 379/28 | No LP | **-** | - | BUV395 |
| 525/50 | 505 LP | **-** | - | BUV496 |
| 740/35 | 690 LP | **-** | - | BUV737 |
| **405 nm** | | **405 nm** | |  |
| 450/50 | No LP | 450/50 | No LP | BV421 |
| 525/50 | 505 LP | 525/50 | 505 LP | BV480, BV510 |
| - | - | 586/15 | 570 LP | - |
| 610/20 | 600 LP | 610/20 | 600 LP | BV605 |
| 670/30 | 635 LP | 660/20 | 630 LP | BV650 |
| 710/50 | 685 LP | 710/50 | 685 LP | BV711 |
| 780/60 | 750 LP | 780/60 | 750 LP | BV786 |
| **488 nm** |  | **488 nm** |  |  |
| 488/10 | No LP | 488/10 | No LP | SSC |
| 530/30 | 505 LP | 530/30 | 505 LP | FITC, BB515 |
| 710/50 | 685 LP | 710/50 | 630 LP | PerCP-Cy5.5, BB700 |
| **561 nm** |  | **561 nm** |  |  |
| 586/15 | No LP | 582/15 | No LP | PE |
| 610/20 | 600 LP | 610/20 | 600 LP | PE-CF594, PE-Vio615 |
| 675/50 | 635 LP | 685/35 | 635 LP | - |
| 780/60 | 750 LP | 780/60 | 750 LP | PE-Cy7, PC7 |
| **640 nm** |  | **640 nm** |  |  |
| 670/30 | No LP | 670/14 | No LP | APC |
| 730/45 | 690 LP | 730/45 | 690 LP | Fixable Viability Stain 700 |
| 780/60 | 750 LP | 780/60 | 750 LP | APC-H7, APC-Cy7 |

**Supplementary Table 4. Target values for 7^th^ peak of rainbow beads in fluorescent channels**

| **Fluorochrome** | **Channel** | Lower (-15%) | **Target MFI** | Upper (+15%) | Recommendation |
| --- | --- | --- | --- | --- | --- |
| **BUV395** | **UV395** | 17,000 | **20,000** | 23,000 | In-house |
| **BUV496** | **UV525** | 23,800 | **28,000** | 32,200 | In-house |
| **BUV737** | **UV737** | 21,2500 | **25,000** | 28,750 | In-house |
| **BV421** | **V450** | 100,452 | **118,178** | 135,905 | EuroFlow |
| **BV480/BV510** | **V525** | 93,871 | **110,436** | 127,002 | EuroFlow |
| **BV605** | **V610** | 47,731 | **56,154** | 64,577 | In-house |
| **BV650** | **V670** | 34,383 | **40,450** | 46,518 | In-house |
| **BV711** | **V710** | 15,079 | **17,740** | 20,401 | In-house |
| **BV786** | **V780** | 1,903 | **2,239** | 2,575 | In-house |
| **FITC/BB515** | **B530** | 28,752 | **33,826** | 38,900 | EuroFlow |
| **PerCP-Cy5.5/ BB700** | **B710** | 66,846 | **78,642** | 90,438 | EuroFlow |
| **PE** | **YG586** | 32,381 | **38,095** | 43,809 | EuroFlow |
| **PE-CF594/**  **PE-Vio615** | **YG610** | 178,500 | **210,000** | 241,500 | In-house |
| **PE-Cy7** | **YG780** | 8,316 | **9,783** | 11,250 | EuroFlow |
| **APC** | **R670** | 158,639 | **186,634** | 214,629 | EuroFlow |
| **AF700** | **R730** | 121,550 | **143,000** | 164,450 | In-house |
| **APC-H7** | **R780** | 64,194 | **75,522** | 86,850 | EuroFlow |
| Spherotech Rainbow Calibration particles (8 peaks) 3.41µm; cat nr. RCP-30-5A, Lot No. EAG01  As per EuroFlow recommendation (Kalina *et al.* 2012) | | | | | |

**Supplementary Figures (n=6)**

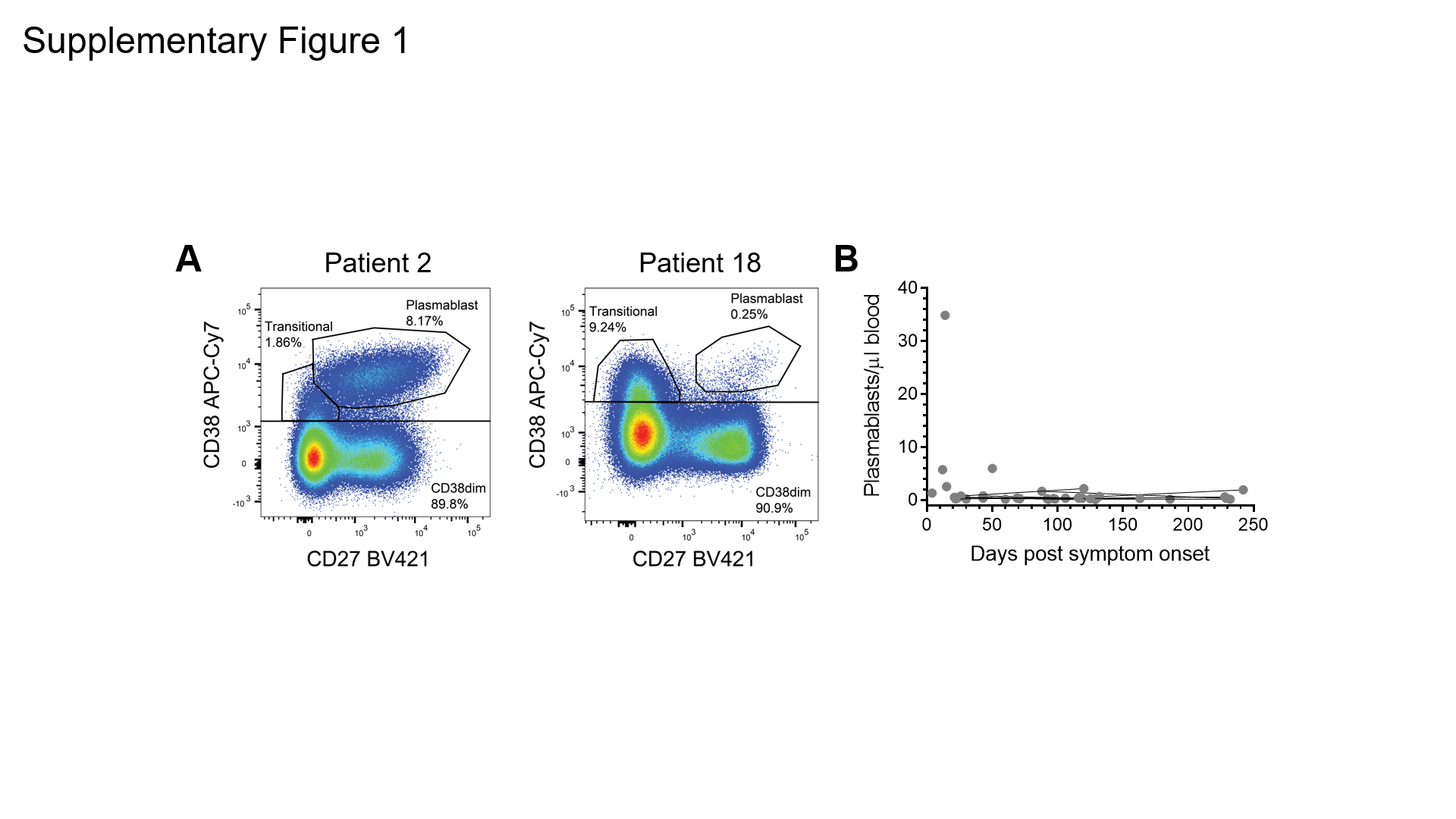
**Supplementary Figure 1. Absolute numbers of plasmablasts in COVID-19 patients**

**A)** Examples of plasmablast populations (CD38^high^CD27^+^) gated within total CD19^+^ B cells of two patients: patient 2, early in SARS-CoV-2 infection and patient 18, in convalescence (**Tables 1 and 2**). **B)** Absolute numbers of plasmablasts in 25 COVID-19 patients of which 11 patients sampled twice. Paired samples are connected with black lines.

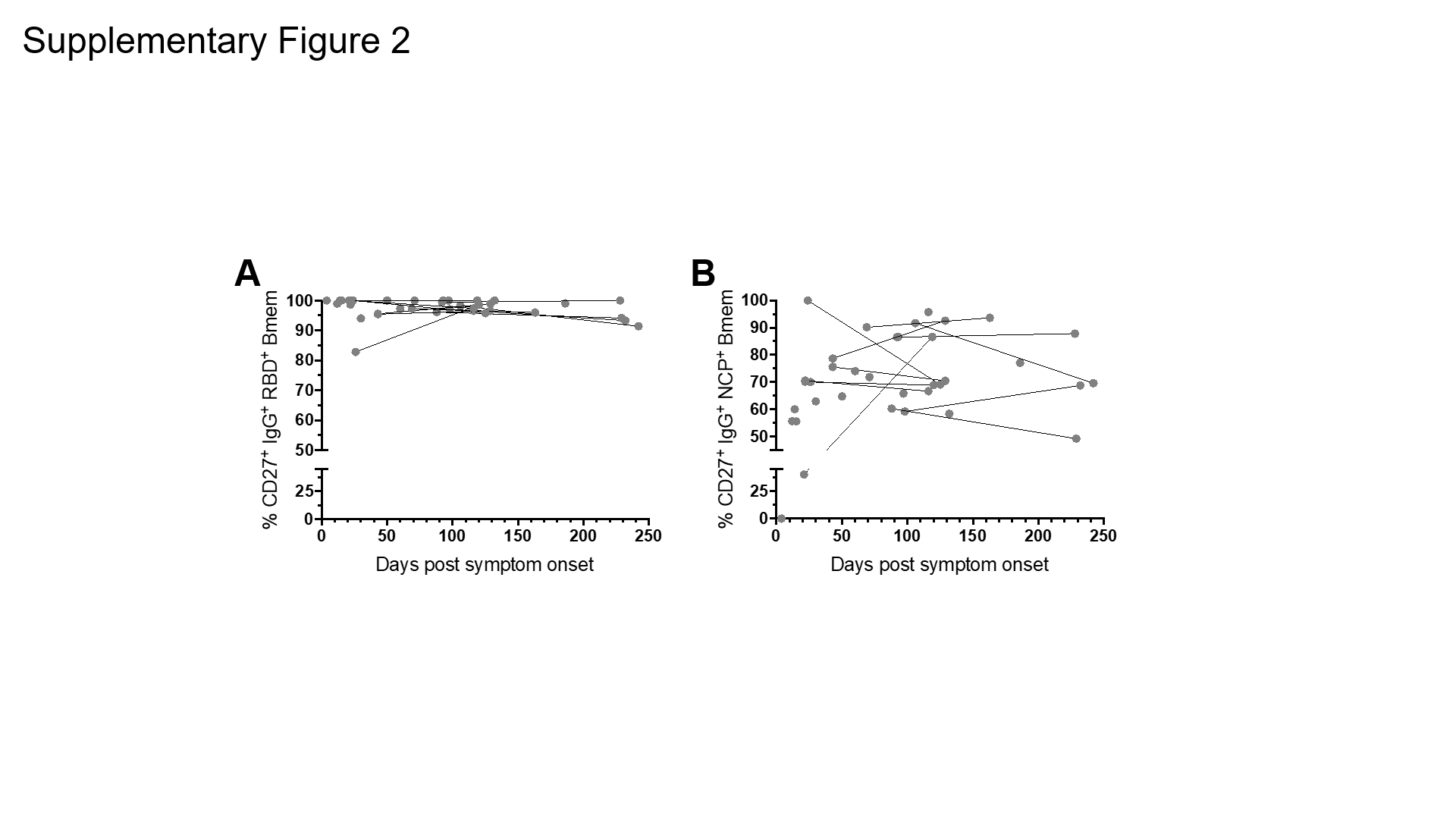

**Supplementary Figure 2. Frequencies of RBD- and NCP-specific IgG^+^ Bmem cells expressing CD27.** Frequencies of **A)** RBD-specific (RBD^+^) and **B)** NCP-specific (NCP^+^) IgG^+^ Bmem cells expressing CD27 versus time since symptom onset for 25 COVID-19 patients, with 11 patients sampled twice. Paired samples are connected with grey lines.

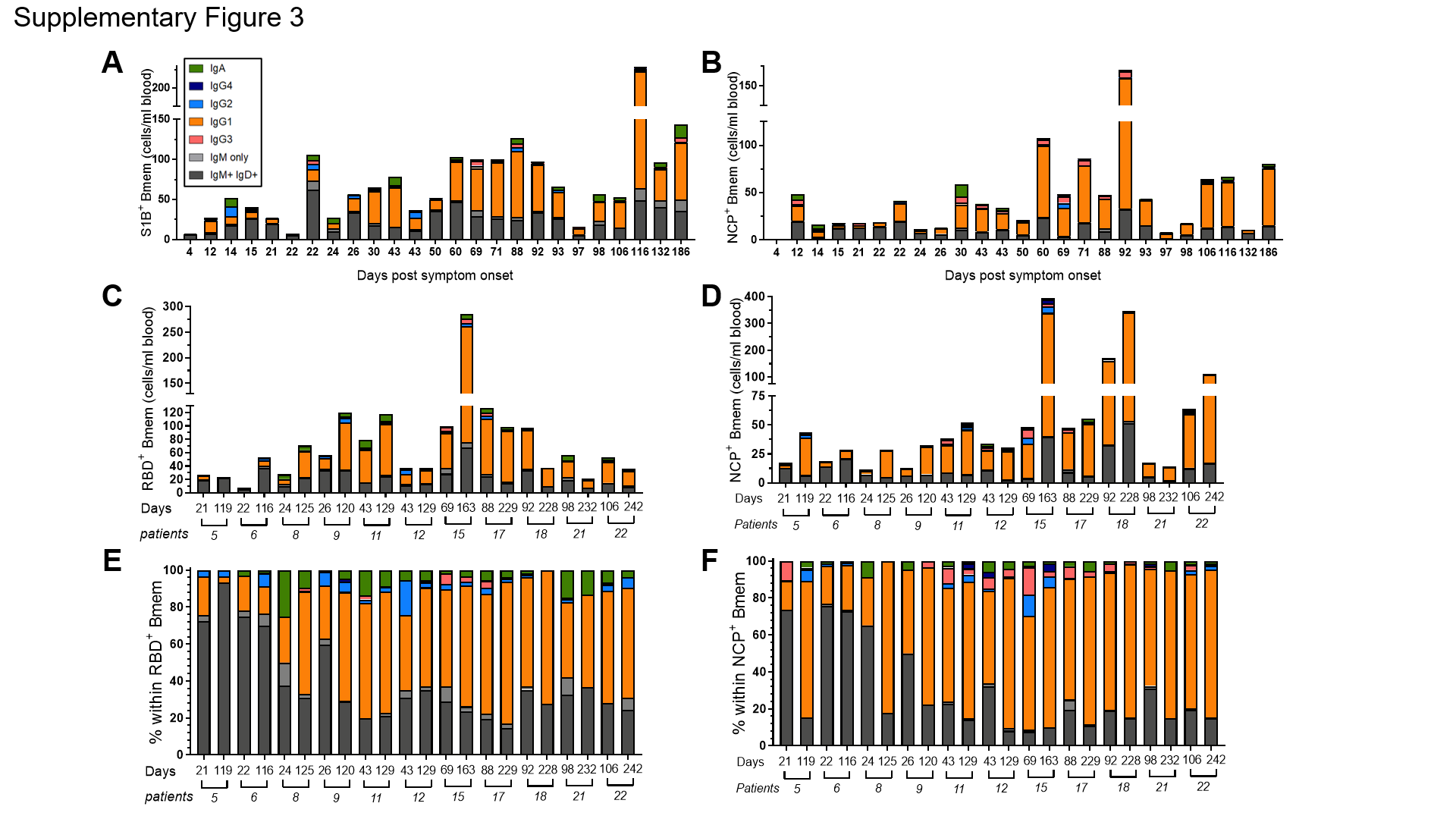
**Supplementary Figure 3. Absolute numbers and frequencies RBD- and NCP-specific Bmem cells expressing distinct Ig isotypes and IgG subclasses.** Absolute numbers of **A)** RBD-specific (RBD^+^) and **B)** NCP-specific (NCP^+^) Bmem cells in all first samples of 25 patients. Absolute numbers of **C)** RBD^+^ and **D)** NCP^+^ Bmem cells from 11 paired samples. Relative distributions of Ig isotype and IgG subclass expression subsets within **E)** RBD^+^ and **F)** NCP^+^ Bmem cells for 11 paired samples. In all panels, patients are ordered by days post-symptom onset.

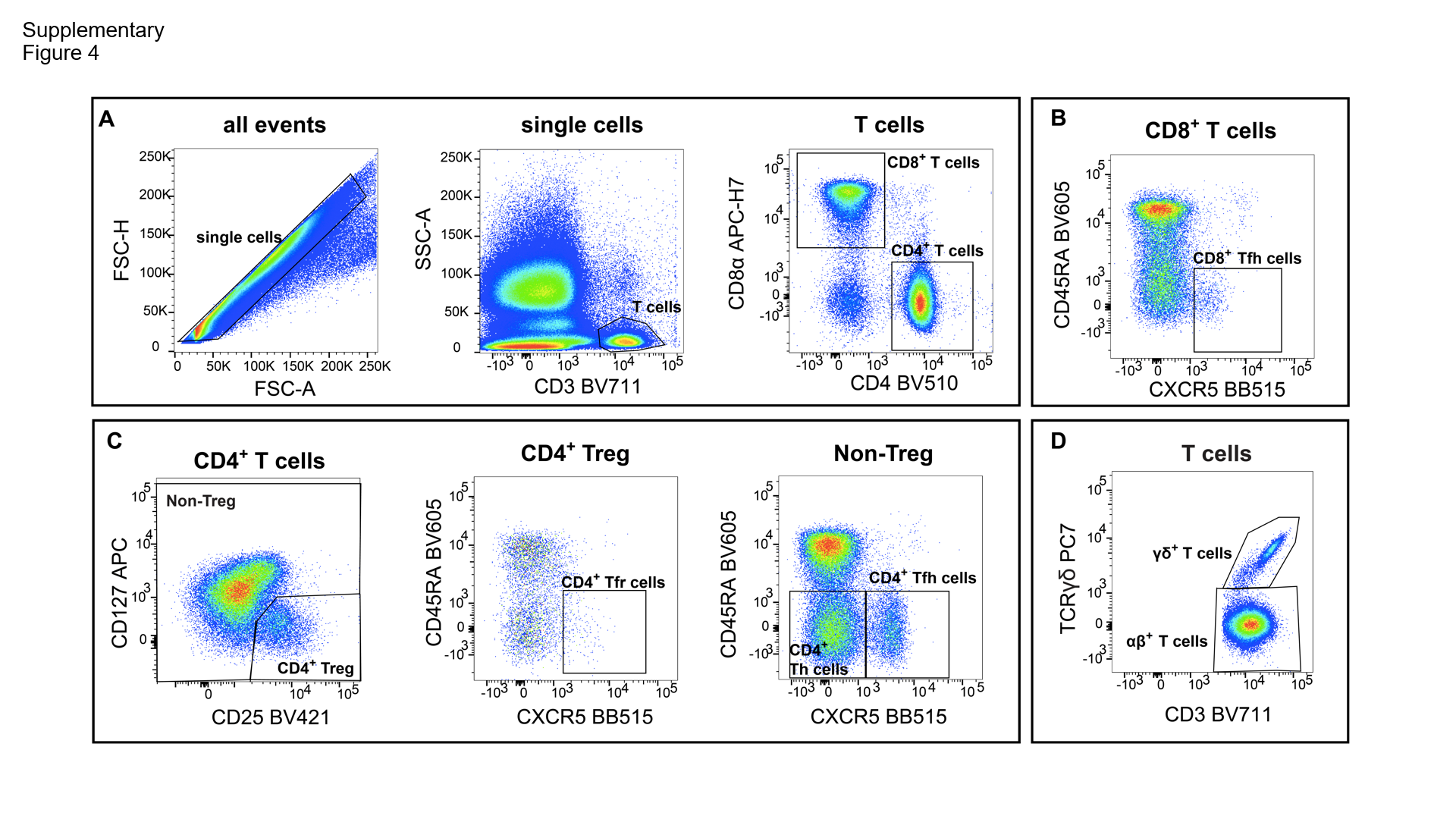

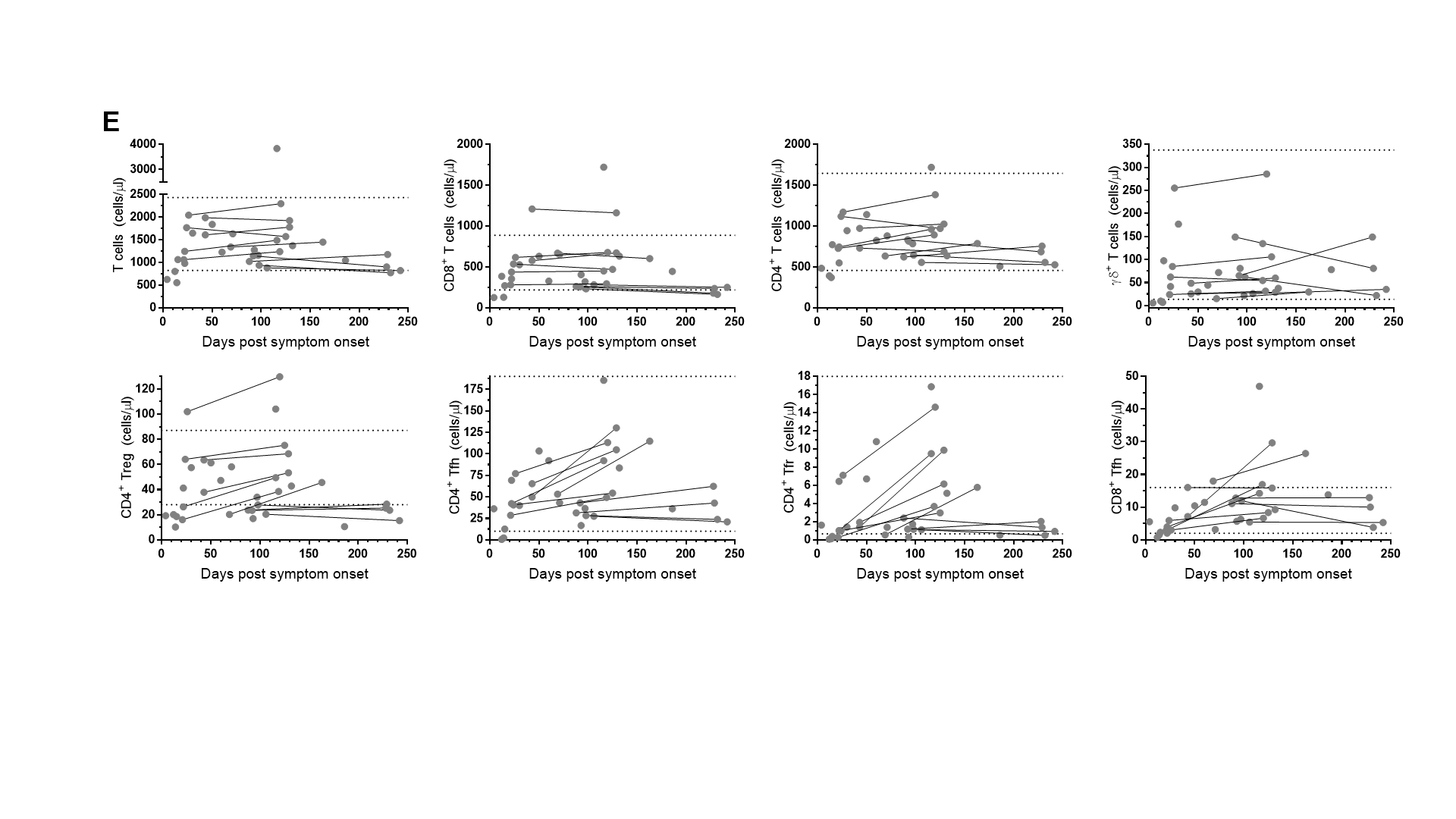
**Supplementary Figure 4. Absolute numbers of T helper cell subsets in COVID-19 patients. A)** Gating strategy to delineate CD3^+^, CD4^+^ and CD8^+^ T cells. **B)** CD8^+^ Tfh gating within total CD8^+^ T cells. **C)** Gating strategies for CD4+ Treg, Tfr and Tfh cells. **D)** Gating strategy for γδ^+^ T cells within the total CD3^+^ population. **E)** Absolute numbers of total T cells, CD8^+^, CD4^+^, γδ^+^, CD4^+^ Treg, CD4^+^ Tfr, CD4^+^Tfh, and CD8^+^ Tfh subsets are plotted versus days post-symptom onset for 25 COVID-19 patients, with seven patients sampled twice. Paired samples are connected with grey lines. Horizontal dotted lines represent the 5th and 95th percentiles of the control group as defined previously (Edwards *et al.* 2019). Statistics were performed using the Wilcoxon matched-pairs signed rank test and the non-parametric Spearman’s rank correlation; * *p* < 0.05.

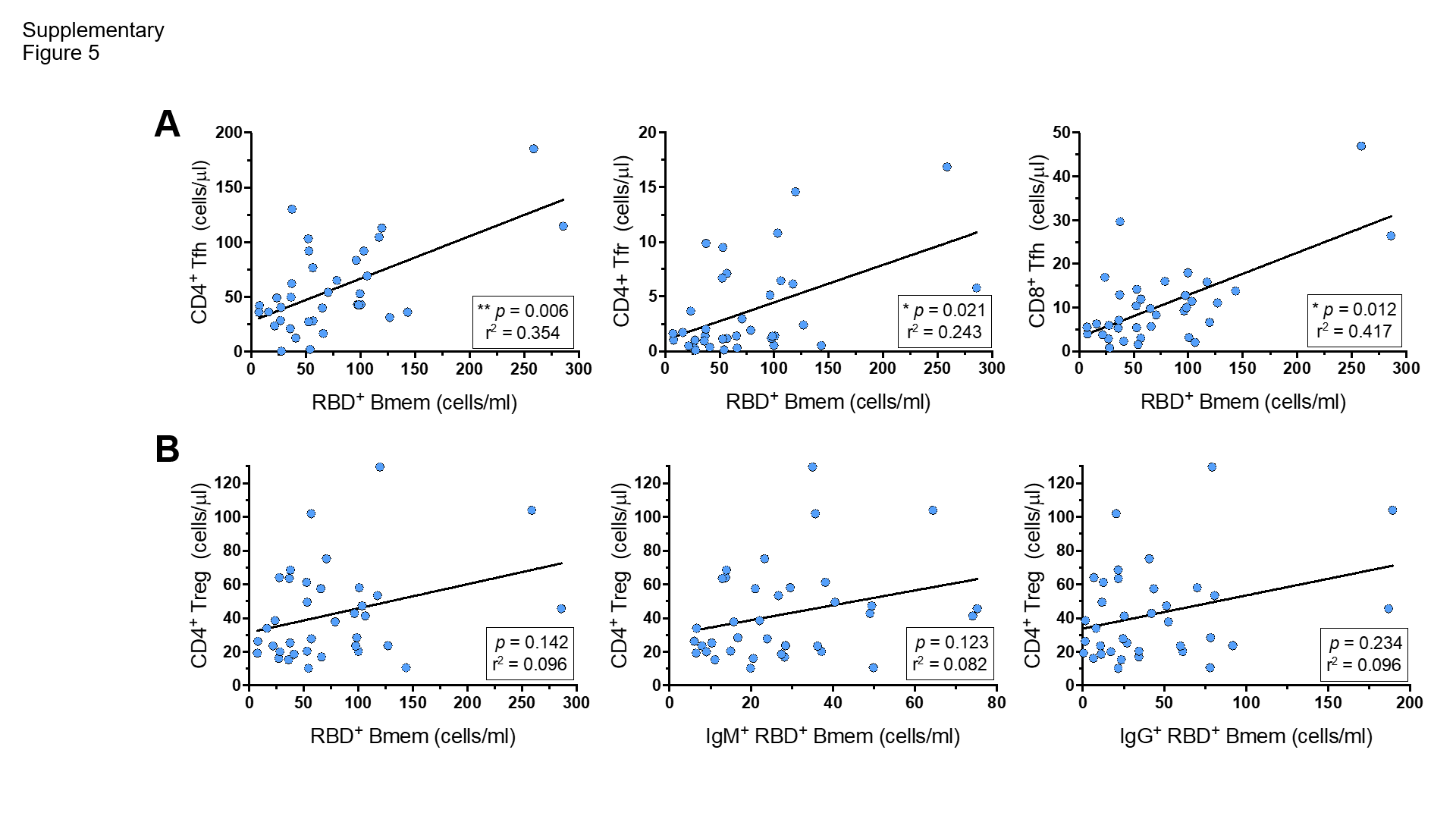
**Supplementary Figure 5. Correlations between absolute numbers of RBD-specific Bmem and T helper cell subsets.**

**A)** Correlations between absolute numbers of RBD-specific (RBD^+^) Bmem cells and CD4^+^ Tfh, CD4^+^ Tfr and CD8^+^ Tfh cells. **B)** Correlations between CD4^+^ Treg cells and RBD^+^ Bmem cells, IgM^+^ Bmem cells and IgG^+^ Bmem cells. For population definitions, see **Supplementary Figure 4**. Trend lines depict linear correlations, statistics were performed using the non-parametric Spearman’s rank correlation; * *p* < 0.05, ** *p* < 0.01.

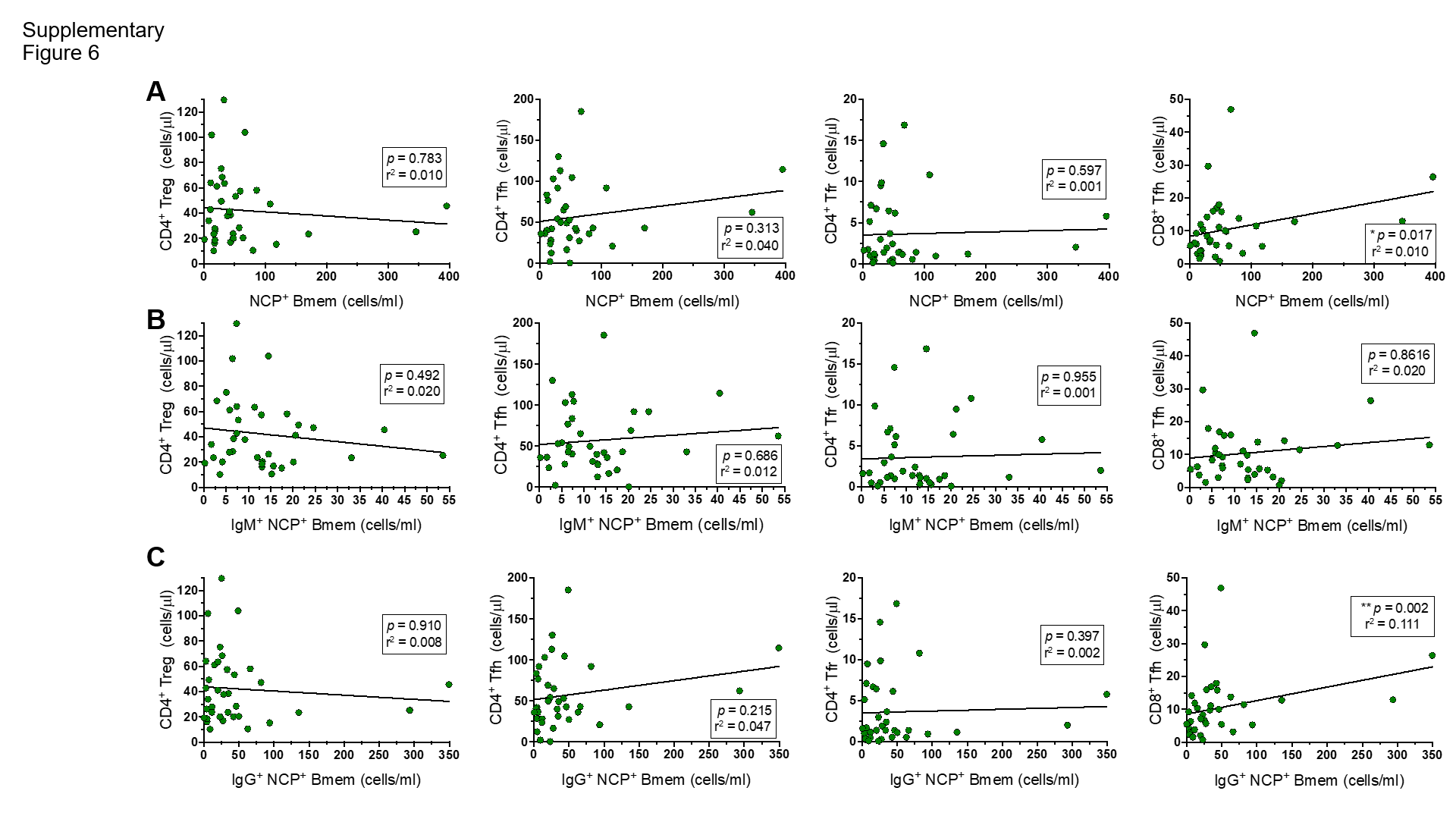
**Supplementary Figure 6. Correlations between absolute numbers of NCP-specific Bmem and T helper cell subsets.** Correlations between CD4^+^ Treg, CD4^+^ Tfh, CD4^+^ Tfr and CD8^+^ Tfh cells and **A)** NCP-specific (NCP^+^) Bmem cells, **B)** NCP^+^ IgM^+^ Bmem cells and **C)** NCP^+^ IgG^+^ Bmem cells. For population definitions, see **Supplementary Figure 4**. Trend lines depict linear correlations, statistics were performed using Spearman’s rank correlation; * *p* < 0.05, ** *p* < 0.01.
